# Multi-trait and multi-ancestry genetic analysis of comorbid lung diseases and traits improves genetic discovery and polygenic risk prediction

**DOI:** 10.1101/2024.08.25.24312558

**Authors:** Yixuan He, Wenhan Lu, Yon Ho Jee, Ying Wang, Kristin Tsuo, David C. Qian, James A. Diao, Hailiang Huang, Chirag J. Patel, Jinyoung Byun, Bogdan Pasaniuc, Elizabeth G. Atkinson, Christopher I. Amos, Matthew Moll, Michael H. Cho, Alicia R. Martin

## Abstract

While respiratory diseases such as COPD and asthma share many risk factors, most studies investigate them in insolation and in predominantly European ancestry populations. Here, we conducted the most powerful multi-trait and -ancestry genetic analysis of respiratory diseases and auxiliary traits to date. Our approach improves the power of genetic discovery across traits and ancestries, identifying 44 novel loci associated with lung function in individuals of East Asian ancestry. Using these results, we developed PRSxtra (cross TRait and Ancestry), a multi-trait and -ancestry polygenic risk score approach that leverages shared components of heritable risk via pleiotropic effects. PRSxtra significantly improved the prediction of asthma, COPD, and lung cancer compared to trait- and ancestry-matched PRS in a multi-ancestry cohort from the All of Us Research Program, especially in diverse populations. PRSxtra identified individuals in the top decile with over four-fold odds of asthma and COPD compared to the first decile. Our results present a new framework for multi-trait and -ancestry studies of respiratory diseases to improve genetic discovery and polygenic prediction.

## Introduction

Respiratory diseases are leading causes of morbidity, mortality, and disability-adjusted life-years globally. Chronic obstructive pulmonary disease (COPD) is the third leading cause of death globally, asthma is the most common chronic disease of childhood, and lung cancer is the leading cause of cancer deaths worldwide^1–3^.

Existing models for predicting the risk of respiratory disease are limited and often do not consider environmental or genetic risk factors beyond smoking. The asthma predictive index (API) determines the likelihood of pediatric asthma mainly based on family history information^4^; and lung cancer and COPD risk are primarily assessed by age and smoking history.

While respiratory diseases such as COPD and lung cancer are strongly influenced by smoking, they are complex diseases that develop due to the influence of many environmental and genetic risk factors. For instance, cumulative non-smoking factors can better predict and stratify COPD risk compared to smoking alone^5^. Genetic factors also play an important role. Family-based studies estimate heritability at 40% to 60% for COPD and asthma^6–8^, respectively, and around 18% for lung cancer^9^, indicating a substantial genetic component.

Previous studies have shown that single-trait polygenic risk scores (PRS), which model the cumulative genetic risk using genome-wide association study (GWAS) summary statistics, can identify and stratify individuals with risk of respiratory diseases ^10,11^. While respiratory diseases share many comorbidities as well as genetic, clinical, and lifestyle risk factors, such as smoking exposure, most PRS to date have been constructed in the context of a single trait and ancestry group without modeling dense genetic correlations between traits and linkage disequilibrium (LD) and allele frequency patterns between ancestries.

To address these limitations, we expanded genetic studies of spirometry, an important diagnostic and management tool for respiratory disease, and developed a novel multi-trait and multi-ancestry PRS approach that integrates genetic correlations between respiratory disease and auxiliary traits and LD patterns across diverse populations. This approach improves the power for genetic discovery across traits and the predictive accuracy of PRS for respiratory diseases, thereby providing a more comprehensive risk assessment tool that can be used in research contexts or potentially integrated into existing clinical models.

Previous efforts have used multi-trait approaches to improve genetic discovery and prediction^12^. For example, multi-trait analysis of GWAS (MTAG) has demonstrated significant improvements in the power of detecting signals for psychiatric disorders, cardiomyopathies, and tobacco and alcohol use, among others ^13–15^. Multi-trait PRS constructed from a weighted sum of multiple cardiometabolic trait scores also predicts heart disease better than the single trait PRS^16,17^. Similarly, lung function PRS from two spirometry measurements–Forced Expiratory Volume in one second (FEV_1_) and the ratio of FEV_1_ to Forced Vital Capacity (FVC)–have been used in predicting asthma and COPD^10,18^. However, the majority of studies still report a single-trait PRS derived in cohorts of primarily European genetic ancestry. This lack of diversity in genomic studies has been shown to greatly reduce the generalizability of prediction models and exacerbate healthcare disparities^19^.

We hypothesize that the joint modeling of multi-trait and -ancestry information at the SNP level will enhance genomic discovery and prediction in respiratory diseases with the largest global disease burden and disparities. Specifically, we jointly model the genetic information of four ancestry groups with population labels and definitions based on training with genetic reference panels, including African (AFR), Admixed American (AMR), East Asian (EAS), and European (EUR)^20,21^. We also model eight strongly correlated traits–COPD, asthma, lung cancer, forced expiratory volume (FEV_1_), forced vital capacity (FVC), FEV_1_/FVC, smoking status, and smoking intensity.

To evaluate the utility of this approach for predicting respiratory diseases and interpreting genetic variant effects across traits and ancestries, we: 1) conducted the largest meta-analysis of lung function in East Asian populations and a comprehensive multi-trait meta-analysis of respiratory disease and related traits to significantly improve genetic discovery; 2) compared shared and distinct genetic architectures and effects by modeling pleiotropy across these traits; 3) developed and validated the PRSxtra (cross TRait and Ancestry) method in the All of Us Research Program, modeling genetic correlations between traits and ancestry-specific LD and allele frequency patterns between populations; and 4) quantified PRSxtra prediction accuracy and case stratification of asthma, COPD, and lung cancer risk across multiethnic populations, especially those traditionally underrepresented in genetic studies, and compared it to single-trait and -ancestry PRS and clinical risk factors.

## Results

We aim to improve the power of genetic discovery and prediction of respiratory diseases through multi-ancestry and multi-trait analyses of eight correlated traits - COPD, asthma, lung cancer, spirometry (FEV_1_, FVC, FEV_1_/FVC), smoking status, and smoking intensity in African (AFR), Admixed (AMR), East Asian (EAS), and European (EUR) ancestry populations. We conducted the largest meta-analysis GWAS of lung function in EAS to date and meta-analyzed additional GWAS from the Global Biobank Meta-analysis Initiative, GWAS & Sequencing Consortium of Alcohol and Nicotine (GSCAN), and multi-population genome-wide meta-analyses of lung function and lung cancer. We then developed the most predictive PRS of asthma, COPD, and lung cancer across populations using the strategy and datasets outlined in **Figure 1**.

**Figure 1.**
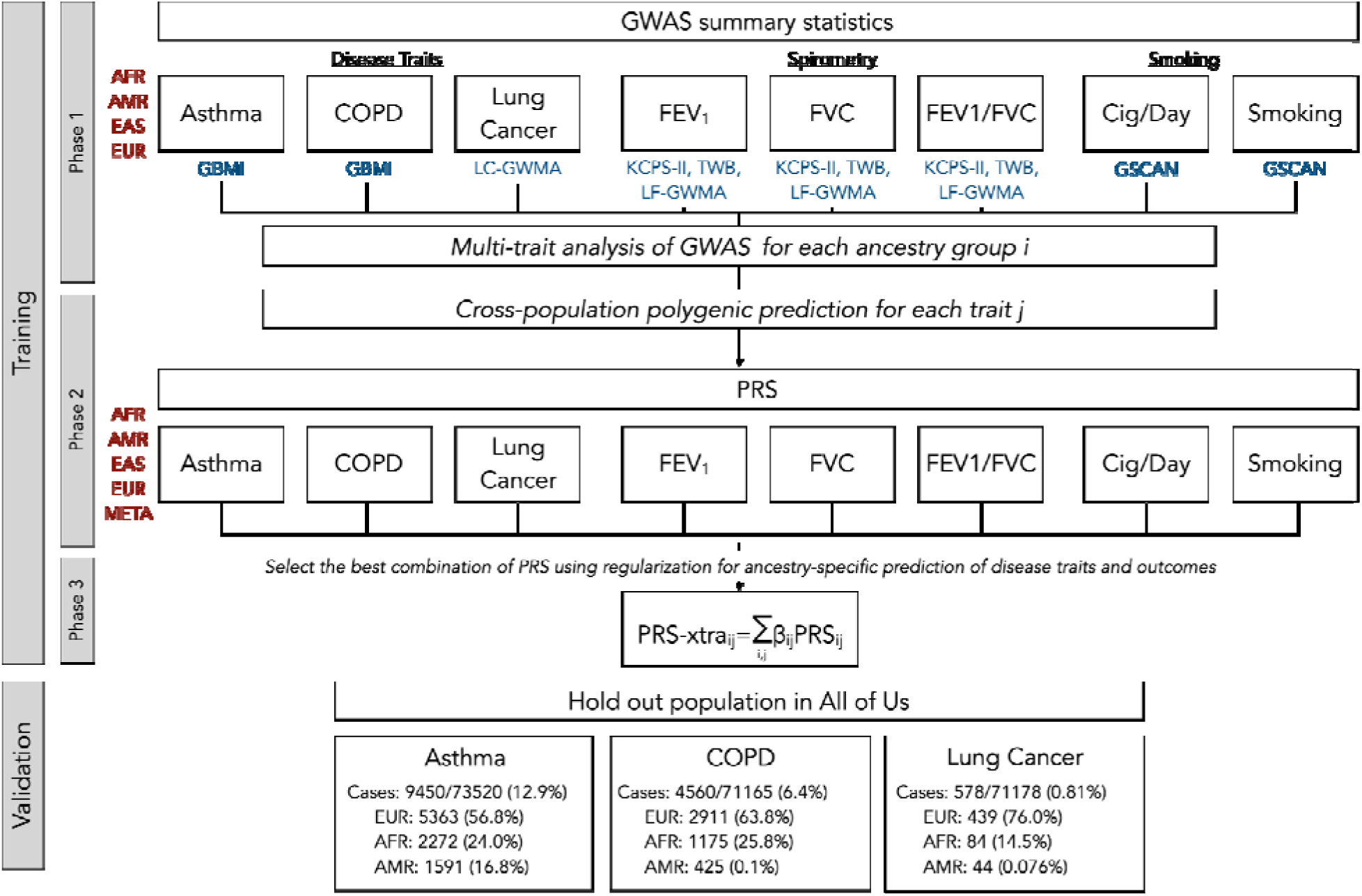
Study design overview. Training PRS models consisted of three phases: Phase 1 = multi-trait analysis, Phase 2 = multi-ancestry analysis, and Phase 3 = regularization to optimally predict traits. We validated PRS in All of Us data for asthma, COPD, and lung cancer case status. GBMI=Global Biobank Meta-analysis Initiative. LC/LF-GWMA=Lung Cancer/Lung Function multipopulation Genome-Wide Meta-Analysis. KCPS-II=Korean Cancer Prevention Study-II. TWB=Taiwan Biobank. GSCAN=GWAS & Sequencing Consortium of Alcohol and Nicotine use. Genetically defined ancestry group labels are based on population reference panels from the 1000 Genomes Project and Human Genome Diversity Project, as follows: AFR=African, AMR=Admixed American, EAS=East Asian, EUR=European, and META=meta-analysis across ancestries.

### Largest genome-wide association study of lung function in the East Asian population identifies novel loci

Spirometry is a pulmonary test that evaluates lung function. As continuous measures of lung function, they are useful traits for studying the heritable basis of respiratory diseases. However, the largest genetic studies of spirometry to date have been Eurocentric. To identify novel loci associated with lung function in understudied populations, we performed the largest GWAS to date of East Asian (EAS) ancestry individuals of FEV_1_, FVC, and FEV_1_/FVC, combining GWAS summary statistics from the Korean Cancer Prevention Study-II (KCPS2) and Taiwanese Biobank (TWB) (**Supplementary Table 1**, Methods)^22–24^. We conducted fixed-effects inverse-variance weighted meta-analysis for the three continuous spirometry measures of lung function across 132,200 total individuals. We applied quality control filters (Methods) that resulted in 8 million unique single-nucleotide polymorphisms (SNPs) for meta-analysis.

The meta-analysis identified 44, 73, and 31 independent loci for FEV_1_, FVC, and FEV_1_/FVC, respectively (**Figure 2, Supplementary Figure 1-3**). Of these, 44 are novel and have not been previously associated with each trait (**Supplementary Table 2**), including 25 novel loci for FVC, 17 for FEV_1_, and 2 for FEV_1_/FVC. Among these novel loci, several were previously associated with height, including rs3782886 in *BRAP* (P=4.20×10^-8^, β=0.0219 with FEV_1_), rs7290267 near *FLJ27365* (P=1.49×10^-9^, β=-0.0272 with FEV_1_), rs724016 in *ZBTB38* (P=1.35×10^-10^, β=-0.0178 with FVC), rs58744877 near *GNAS* (P=2.71×10^-14^, β=-0.033 with FVC), rs3176466 near *CDKN2C* (P=4.59×10^-8^, β=0.0236 with FVC), rs28839214 near *RP11-361D14.2* (P=1.91×10^-13^, β=0.0197 with FVC), rs149580940 in *FIBIN* (P=7.89×10^-10^, β=0.0449 with FVC)^25–27^. rs3782886 has also shown previous associations with other traits including alcohol use disorder, high-density lipoprotein, and type 2 diabetes^28–30^. rs724016 has also been shown to be associated with Crohn’s disease^31^, and rs150971595 near *GABBR1* (P=1.49×10^-8^, β=-0.0307 with FVC) associated with total cholesterol level and triglyceride levels^32^. Previous reported associations for each locus can be found in **Supplementary Table 3**.

**Figure 2.**
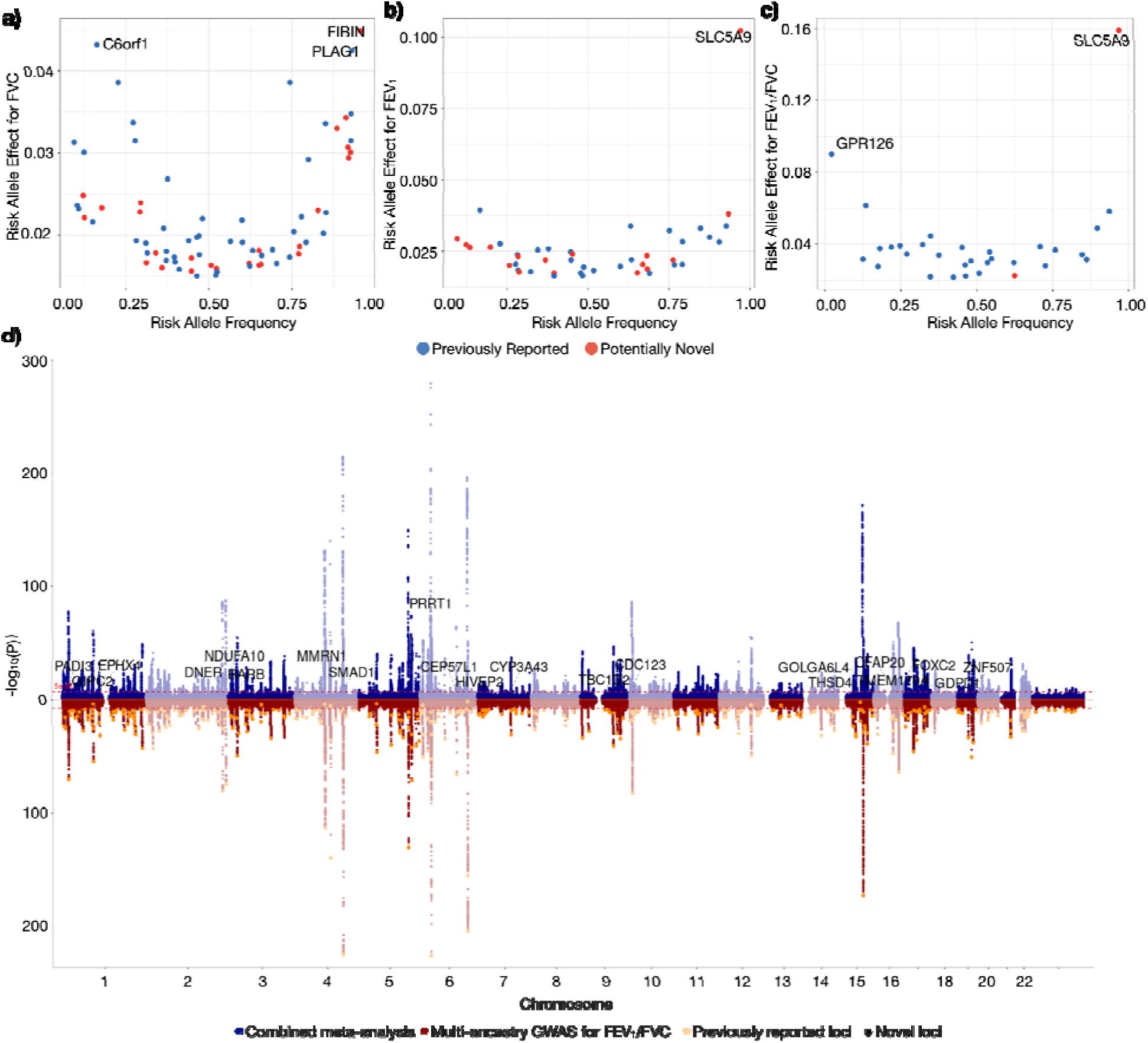
Meta-analysis results of spirometry in East Asian cohorts. Frequency and effect size of risk alleles of 44, 73, and 31 index variants for FVC (a), FEV_1_ (b), and FEV_1_/FVC (c). Novel associations are highlighted in red. Genes with large effect sizes are highlighted. d) Results of our combined EAS meta-analysis of FEV_1_/FVC with the largest multi-ancestry GWAS to date. The Manhattan plot of the previous multi-ancestry GWAS is shown in red on the bottom, with yellow dots representing previously identified loci associated with FEV_1_/FVC. The Manhattan plot of the meta-analyzed results of the previous study with our new EAS meta-analysis is shown in blue on the top, with new loci annotated and indicated with black stars. Gene names indicate the nearest gene to novel loci.

We combined the results of our EAS meta-analysis of lung function with the largest multi-ancestry GWAS of lung function to date^33^. Incorporating results from KCPS2 and TWB resulted in 519, 526, and 476 total loci discovered for FEV_1_, FVC, and FEV_1_/FVC, respectively, of which 131, 163, and 149 were not originally reported to be associated with each trait (**Figure 2, Supplementary Figure 4-5**). Significantly associated loci are reported in **Supplementary Table 4-6**.

### Pervasive pleiotropic effects across respiratory traits and diseases inform shared components of heritable risk

Measures of lung function are known clinical risk factors for respiratory diseases such as chronic obstructive pulmonary disease (COPD), asthma, and lung cancer. Using the largest multi-ancestry GWAS summary statistics to date, we found that lung function (FEV_1_, FVC, FEV_1_/FVC), smoking behavior (smoking status and cigarettes/day), and respiratory diseases (asthma, COPD, and lung cancer) were all genetically significantly correlated with each other (P<0.05) with the exception of FVC and lung cancer (**Figure 3a**).

**Figure 3:**
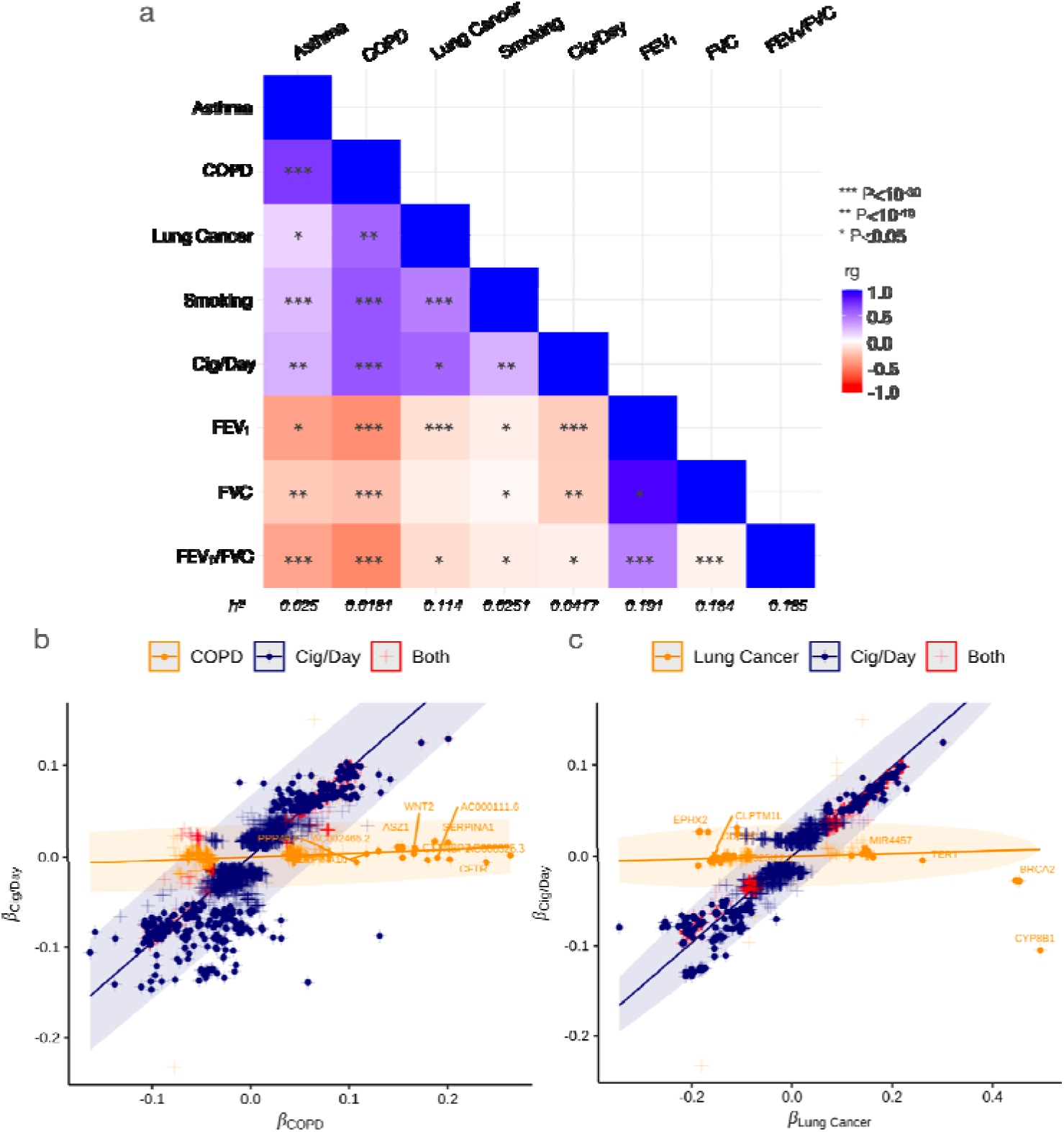
Shared and distinct heritable components inform the molecular basis of trait differences. a) Genetic correlations between traits (top) and heritability estimates (bottom) analyzed in this study based on S-LDSC correlations. Colors indicate magnitude of correlation and stars indicate degree of significance. Comparison of effect sizes of variants from GWAS on EUR samples for (b) COPD vs. Cigarettes smoked per day (Cig/Day) and (c) Lung cancer vs. Cig/Day. Effect sizes of variants on diseases (COPD and lung cancer) are on the x-axis and effect sizes of variants on Cig/Day are on the y-axis. Each cross represents a variant significantly associated (p< 5x10^-8^) with at least one of the corresponding pair of traits, colored by their significance status: variants significantly associated with both traits (red), variants significantly associated with disease (COPD or Lung cancer) only (orange), and variants only significantly associated with Cig/Day (navy). Solid dots are variants confidently identified to have predominant effect on one of the two traits by a Bayesian classifier (linemodels) in shared variants analysis with posterior probability >99%: variants predominantly associated with disease (orange), and variants predominantly associated with Cig/Day (navy). The colored ellipse range indicates the 95% probability regions of the fitted bivariate effect size distributions with each class. Nearest gene names are labeled on disease-specific variants with the highest posterior probability for each gene.

To examine the distinct and shared variants among respiratory diseases and auxiliary traits, we first evaluated the consistency of lead SNPs between the traits. Many variants demonstrated pleiotropic effects associated with multiple traits. In particular, we explored the shared and distinct genetic etiology among the three respiratory disease traits (asthma, COPD, and lung cancer), risk factors (smoking status, smoking intensity), and a measure of lung function (FEV_1_/FVC). For each trait pair, we selected variants significantly associated with either or both of the traits. We computed the linear slopes of variant effect sizes within each category using an expectation-maximization (EM) algorithm, which typically revealed more than one linear trend and divergent slopes (Methods, **Figure 3**, **Supplementary Figure 6-8**). For example, genetic variants associated with the amount and frequency of smoking have mostly distinct effects from asthma. This suggests that there is no uniform explanation of the genetic mechanisms of the two traits across those variants.

We then implemented a Bayesian algorithm, linemodels^34^, to classify the variant effect sizes into confident associations with trait 1 or trait 2, as smoking is a causal risk factor for both diseases. A model that fits three lines, including both traits, did not significantly improve the fit but is shown in **Supplementary Figures 9-11**. We used ancestry-specific (AFR, EAS, EUR) as well as meta-analyzed multi-ancestry GWAS results to identify variants having confident associations with a posterior probability above 0.99. We found 15 variants that were confidently associated with COPD and 88 with lung cancer, which are distinct from those associated with smoking intensity, as measured by Cigarettes/Day, in EUR. We identified 690 and 486 variants, respectively, confidently associated with only smoking intensity in each analysis. (**Supplementary Table 7-8**).

When we further investigated variants with predominant effects on lung cancer across EAS, EUR, and the multi-ancestry meta-analysis GWAS results, we identified 6 variants from *TERT* appearing in all three groups and 11 variants significant only in the meta-analysis results (**Supplementary Figure 12-13)**. These include rs11783093 and rs73229093 from *GULOP*, rs11778371 from *CHRNA2*, 4 variants from *CLPTM1L* and 5 other variants within or near *TERT*.

### PRSxtra jointly models multi-ancestry and multi-trait effects to predict diseases and exacerbations

To develop PRSxtra, we combined the results of our new GWAS meta-analysis of lung function in EAS with the largest and most diverse GWAS of COPD, asthma, lung cancer, and smoking (**Figure 1**) to conduct ancestry-specific multi-trait GWAS (MTAG)^12^. We then used PRS-CSx to jointly model trait-specific MTAG results across ancestry groups to derive candidate scores^35,36^ to include in the final PRSxtra.

First, MTAG resulted in a greater number of significant associations and loci discovered across traits (**Supplementary Table 9**). AMR and EUR ancestry-specific MTAG analysis resulted in the largest gains. For example, FVC in AMR and COPD in EUR cohorts had the greatest proportional increases in the number of lead SNPs discovered, from 2 to 34 (17-fold) and 27 to 441 (16-fold), respectively. In total, MTAG identified 609 more lead SNPs across all traits and ancestry groups. We assessed the gain in power for each run of MTAG by the increase in mean χ^2^ statistic, where all but five GWAS (all of which were ancestry-specific spirometry GWAS) increased in power (**Supplementary Table 9**).

Next, we aggregated the summary statistics from MTAG to derive multi-trait and multi-ancestry polygenic risk scores. For each trait, we used PRS-CSx to jointly model trait-specific MTAG summary statistics across ancestry groups to derive a total of 39 candidate scores for individuals in the All of Us Research Program (AoU), a longitudinal cohort study continuously enrolling adults in the US with genotype, self-reported, and linked health record data. The program places a strong emphasis on including diverse populations that have traditionally been underrepresented in biomedical research. The individuals from AoU are independent from the cohorts in which the GWAS was obtained. For each trait, we randomly split participants who passed quality control into 70% for training and 30% for validation (**Figure 1**, Methods, **Supplementary Table 10-13**). We then used ridge regression to calculate linear combinations of the scores in the training set (**Supplementary Table 14-16, Supplementary Figure 14-16**).

We evaluated the performance of PRSxtra in the held-out validation cohort for predicting asthma (N_cases_=9450, N_controls_=64070), COPD (N_cases_=4560, N_controls_=66605), and lung cancer (N_cases_=578, N_controls_=70600) compared to a single ancestry- and trait-matched PRS (**Figure 1, Supplementary Table 10**). The validation cohort includes individuals of AFR, AMR, EAS, EUR, MID, and SAS genetic ancestries. In this multi-ancestry validation cohort, ancestry- and trait-matched PRS and PRSxtra were significantly correlated with each other (P<0.0001), with r=0.444, r=0.185, and r=0.120 for asthma, COPD, and lung cancer, respectively.

PRSxtra alone predicted asthma, COPD, and lung cancer more accurately than the trait- and ancestry-matched PRS alone (P<0.0001) in the multi-ancestry validation cohort (**Supplementary Table 17-19**). For asthma, the AUC improved from 0.543 (95% CI=[0.537, 0.549]) to 0.563 (95% CI=[0.557, 0.569]) using PRSxtra versus PRS. For COPD, AUC improved from 0.540 (95% CI=[0.532, 0.549]) to 0.589 (95% CI=[0.581, 0.597]). For lung cancer, AUC improved from 0.539 (95% CI=[0.516, 0.561]) to 0.592 (95% CI=[0.569, 0.616]) (**Figure 3**).

Among individuals with each disease, the median percentile of PRSxtra was much higher than PRS (**Supplementary Table 20**). We observed the largest difference between scores in lung cancer, where cases had a median PRSxtra in the 64th percentile compared to the 57th percentile for PRS. Controls had a median PRSxtra and PRS both in the 50th percentile. The AMR population had the largest improvement in disease prediction between PRS and PRSxtra. For asthma, AUC improved from 0.509 (95% CI=[0.493, 0.524]) to 0.630 (95% CI=[0.616, 0.645]). For COPD, AUC improved from 0.540 (95% CI=[0.532, 0.549]) to 0.589 (95% CI=[0.581, 0.597]). These differences could reflect the increased EAS GWAS power for spirometry and the relatively low genetic divergence between EAS and AMR groups. However, these new ancestry-specific GWAS do not explain the most phenotypic variation when predicting these traits. Specifically, we further assessed the robustness of PRSxtra by conducting leave-one-out analyses for predicting COPD and asthma in the AMR cohort, where there were the largest improvements in prediction. Removing any single score did not significantly change the prediction ability of PRSxtra. For example, removing the EUR lung cancer candidate PRS resulted in the largest decrease in AUC of -0.0175 for predicting COPD, and removing EUR FVC candidate PRS resulted in the largest decrease in AUC of -0.000421 for predicting asthma.

PRSxtra improved overall stratification in the multi-ancestry validation cohort compared to PRS. In the bottom and top deciles of PRSxtra, asthma prevalence was 8.4% and 16.8% (+8.4%), respectively, versus 9.0% and 14.1% (+5.1%) for PRS. For COPD, the prevalence in the bottom and top deciles of PRSxtra was 2.8% and 9.1% (+6.3%), respectively, compared to 4.6% and 7.6% for PRS (+3.0%). Lung cancer prevalence in the bottom and top deciles of PRSxtra was 0.5% and 1.4% (+0.9%), respectively, compared to 0.46% to 0.90% (+0.44%) for PRS (**Supplementary Figure 17, Supplementary Table 21-23**). PRSxtra was also better at identifying individuals with the highest risk of disease. For example, individuals in the top decile of PRSxtra had 3.5 times the odds of COPD compared to those in the first decile, while PRS had 1.7 times the odds. The improvement was most apparent in the AMR population for asthma and COPD. Individuals in the top decile of PRSxtra had 4-fold odds of asthma and COPD compared to the first decile, whereas the top decile of PRS had almost the same odds of asthma and COPD compared to the first decile (**Supplementary Figure 17, Supplementary Table 24-26**).

We compared the performance of PRS and PRSxtra alone to a joint model of the strongest clinical risk factors for each disease, i.e. family history of asthma, and smoking for COPD and lung cancer. Clinical risk factors generally provided the largest improvement in performance from sex and age. In the multivariable model, PRSxtra remained significantly better than PRS for predicting asthma in the multi-ancestry validation cohort and for predicting asthma and COPD in the AMR subgroup. For lung cancer, the risk scores do not add additional information to the multivariable model of sex, age, and smoking status (**Figure 4, Supplementary Table 17-19**).

**Figure 4.**
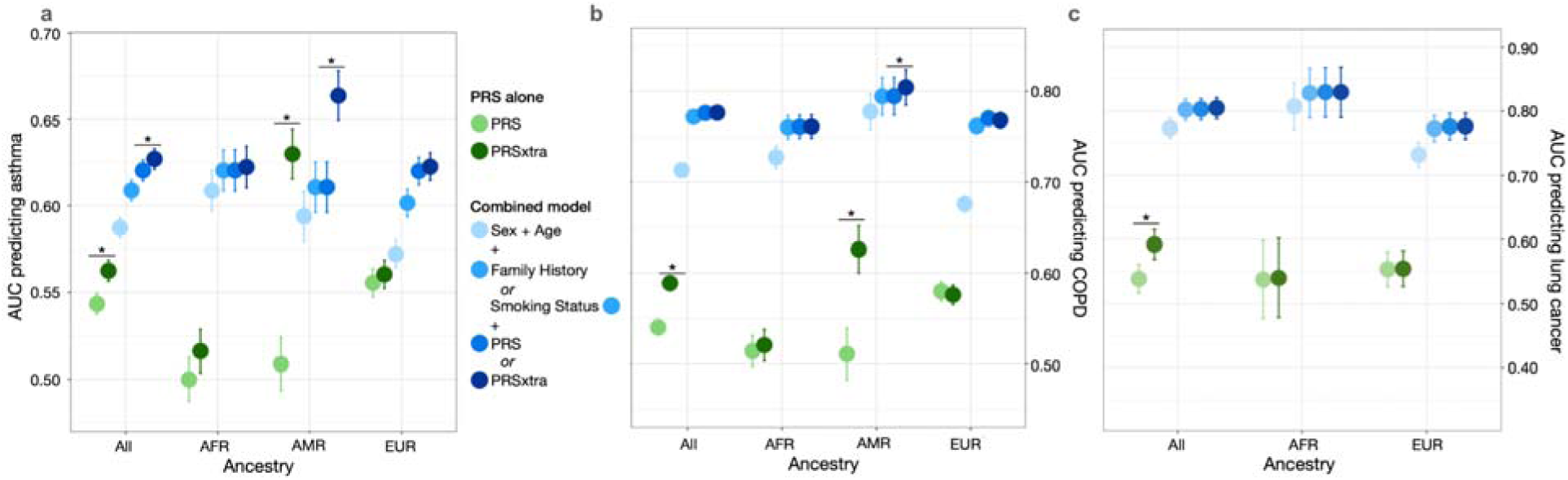
PRSxtra significantly improves prediction for several respiratory diseases compared to PRS and clinical risk factors. Diseases include: asthma (a), COPD (b), and lung cancer (c) in the full multi-ancestry held-out validation set and in ancestry-specific subgroups. Green dots indicate the performance of PRS or PRSxtra alone to predict diseases, blue dots represent the performance of the combined model with incremental variables added from sex and age, family history (for asthma) or smoking status (for COPD and lung cancer), and PRS or PRSxtra in gradually darker shades. The full validation set included individuals of AFR, AMR, EAS, EUR, MID, and SAS predicted genetic ancestry background.

To investigate performance in clinical subgroups, we tested the association of PRSxtra with COPD and lung cancer in subgroups of never and ever smokers, and of asthma in individuals with and without family history. We found similar performance between subgroups (**Supplementary Table 17-19, Supplementary Figure 18-20**). PRSxtra was positively associated with COPD and asthma exacerbations. PRSxtra predicted COPD exacerbation significantly better than PRS in the validation cohort, with AUC increasing from 0.572 (95% CI=[0.558-0.586]) to 0.600 (0.587-0.614), However, PRSxtra did not significantly improve prediction of asthma exacerbation compared to PRS (**Supplementary Table 27**).

## Discussion

In this study, we conducted the most powerful multi-trait and multi-ancestry genetic analysis of respiratory diseases and comorbid traits to date. Our novel study framework contrasts with traditional genetic studies which typically analyze a single trait within a single population, thereby overlooking the genetic correlations between traits and LD patterns across diverse populations.

Our innovative approach, which iteratively models genetic correlations across traits and ancestry groups, significantly increases the power to detect novel genetic associations and enhances disease prediction and risk stratification, especially in non-European ancestry populations. Applying MTAG resulted in a greater number of significant associations and loci discovered across traits, with the largest gains in AMR and EUR ancestry-specific analyses. We also conducted the largest meta-analysis of lung function to date in EAS to identify 44 novel loci associated with lung function. Of these, several are associated with lipid levels such as high-density lipoprotein, total cholesterol level and triglyceride levels^32^, as well as with diseases such as type 2 diabetes and Crohn’s Disease, which supports previously documented relationships between lung dysfunction and disease risk and progression ^37–39^.

Given that smoking is the leading risk factor for COPD and lung cancer, we further investigated the distinct and shared variants between smoking intensity, COPD, and lung cancer. We identified variants that demonstrated clear vertical pleiotropy and had influences on smoking and COPD as well as lung cancer. We also identified variants that distinctly influence disease but not smoking; these variants were confidently associated with COPD and lung cancer but were distinct from those associated with smoking intensity. Among variants confidently associated with COPD but not smoking was the *CFTR* Phe508del mutation rs113993960, a known pathogenic variant for cystic fibrosis^40,41^. rs11571833, a rare variant of *BRCA2*-K3326X, was confidently associated with lung cancer but not smoking and has been shown to have large-effect genome-wide associations for squamous lung cancer (odds ratio=2.47, P=4.74×10^−20^)^42^. Furthermore, across EAS, EUR, and the multi-ancestry meta-analysis GWAS results, there were 6 variants from *TERT* appearing in all three groups and 11 variants in *GULOP, CHRNA2,and CLPTM1L* significant only in the meta-analysis results. *TERT*, *CHRNA2*, and *CLPTM1L*have been shown to have a role in respiratory function: 1) *TERT* mitigates oxidative stress and chronic inflammation, which are essential factors in the disease progression of COPD. It also prevents premature cellular aging and apoptosis in lung epithelial cells to sustain respiratory function. Polymorphisms in *TERT* have previously been associated with pulmonary fibrosis^43^ as well as COPD risk in an EAS population^44^. There is also evidence of strong gene-environment interaction between smoking and telomerase mutations, where carriers who do not smoke predominately develop pulmonary fibrosis, while smokers are also at risk for developing emphysema (alone or in combination with pulmonary fibrosis)^43^. 2) *CHRNA2*, which encodes a subunit of nicotinic acetylcholine receptors, plays a role in respiratory behavior and function by influencing nicotine addiction and smoking habits^45,46^., but has a smaller effect than the *CHRNA3-CHRNA5* cluster. While *CHRNA2* affects respiratory function by modulating the neural pathways involved in nicotine response, the *CHRNA3-CHRNA5* cluster has a stronger and more consistently observed association with Cigarettes/Day^47,48^. 3) *CLPTM1L* is a mitochondrian membrane protein whose overexpression in cisplatin-sensitive cells causes apoptosis^49^. The *CLPTM1L* protein is more highly expressed in lung cancer and plays a protumorigenic role critical for lung cancers driven by mutations in K-Ras^49–51^.

In our investigation of pleiotropic effects across pairs of traits, we applied *linemodels* to dissect the pleiotropic effect of variants. Using this method, we identified unique genetic features of lung cancer and COPD not mediated by one of their leading risk factors - smoking. The ‘linemodels’ algorithm, tailored for pleiotropy dissection, uses a Bayesian framework to probabilistically cluster genetic variants based on their linear relationships in effect sizes across two outcomes. This method optimizes model parameters using the EM algorithm and Gibbs sampling, accommodating correlated estimators due to sample overlaps. In contrast, techniques like Non-negative Matrix Factorization (NMF) decompose data into non-negative factors to capture latent structures without modeling direct linear relationships, highlighting the unique focus of linemodels on linear effect size clustering in genetic studies.

We then developed a cross-trait and -ancestry score, PRSxtra, to model the genetic correlations between related traits and ancestry-specific LD and allele frequency patterns between populations. We derived and validated our score in the multi-ancestry All of Us cohort. Given that one of the strongest genetic risk factors for COPD and emphysema is the alpha-1 antitrypsin deficiency due to rare variants in *SERPINA1*, we excluded individuals with homozygous polymorphism of *SERPINA1* from our study^52,53^. While PRSxtra and the trait- and ancestry-matched PRS are significantly correlated, PRSxtra demonstrates significantly better prediction and stratification for respiratory diseases and disease exacerbations. Previous studies have shown the utility of integrating large and diverse sources of data to construct PRS. For example, GPSmult was calculated by weighting the sum of multiple cardiometabolic trait components, which predicts heart disease better than the single trait PRS^16^. Similarly, PRS derived from two spirometry measurements predict asthma and COPD^10^. Recent developments such as PRSmix and BridgePRS jointly model summary information from multiple traits and ancestry groups at the score level but do not account for genetic correlations across traits or model differences in LD across ancestries. PRSxtra, however, shares information across traits and ancestries at the SNP level, refining putatively causal loci and improving PRS accuracy. We demonstrated the robustness of PRSxtra in a leave-one-out analysis, which showed minimal changes in predictive ability when candidate PRS were excluded. The enhanced predictive capacity of PRSxtra was particularly pronounced in the AMR population, which is not primarily explained by the candidate PRS from the new EAS spirometry GWAS. One potential explanation could be due to the phenotypic heterogeneity of asthma and COPD among Hispanic populations^54–56^. For example, asthma, and COPD are more prevalent in individuals of Puerto Rican heritage than among other Hispanics.

Our study does have limitations. We relied on phenotype definitions based on ICD codes and self-reported data, which can be imprecise. For example, definitions of COPD using common ICD-9 codes have been previously shown to misclassify patients compared to combining them with pharmacy data^57^. Therefore, the performance of PRSxtra may differ for COPD diagnosed by spirometry or by medical diagnostic codes. While we aimed to mitigate this effect by combining multiple ICD-9 and ICD-10 codes to define disease cases, it is unclear how adding spirometry would change disease definitions in AoU due to the lack of spirometry measures availability for all participants. A patient with respiratory symptoms and is a smoker may also be more likely to be labeled as COPD by physicians. We were also limited by the study design of existing GWAS. While traditional GWAS of FEV_1_, FVC, and FEV_1_/FVC are adjusted for smoking status, our meta-analysis of lung function in EAS was not due to the limitation of data availability. Our measures were also not explicitly post-bronchodilator lung function measures, although previous work demonstrated little impact of pre- versus post-bronchodilator definitions when predicting COPD^58,59^. Additionally, we used ridge regression, which assumes a linear association of the candidate PRS. It is unclear how other non-linear methods of combining PRS will compare. Our study sample is shaped by the All of Us Research Program’s cohort creation process, which relies on partnerships with universities, research centers, volunteers, and community engagement^60^. While All of Us is diverse, there are demographic sampling biases that may not be representative of the general population^61^. Our results demonstrate significant improvements in a held-out cohort of diverse AoU participants, but they should be validated in an independent cohort.

In summary, we conducted the largest multi-trait and multi-ancestry genetic analysis of respiratory diseases and auxiliary traits to discover numerous novel genetic signals. We propose PRSxtra as a method to model genetic correlation across traits and LD differences between ancestry groups, significantly improving disease prediction and stratification for asthma, COPD, and lung cancer. PRSxtra has the potential to reduce the disparities in risk stratification between populations for survival and outcome, as well as to advance more equitable and generalizable prediction models for respiratory diseases.

## Methods

### Meta-analysis of lung function across East Asian populations

We performed a fixed-effects meta-analysis with inverse variance weighting, as implemented in METAL v2001-03-25 software, for FEV_1_, FVC, and FEV_1_/FVC across two East Asian cohorts using published summary statistics from Korean Cancer Prevention Study-II (KCPS2)^23,24^, Taiwanese Biobank (TWB)^22,32^. The details of their study design have been described previously. In summary, the Korean Cancer Prevention Study-II Biobank (KCPS2) is a prospective cohort study based in Korea of 153,950 subjects with genotype data and phenotype measurements between 2004 and 2013. The Taiwanese Biobank is a prospective cohort study of the Taiwanese population with 149,894 participants between the ages of 30-70 years old at recruitment (as of April 2021). FEV_1_ was defined as the total air blown between zero and 1 second (measured in Liters). FVC was defined as the vital capacity during forced expiration (measured in Liters). In KCPS2, FEV_1_ and FVC were measured using pulmonary/metabolic systems Vmax 20, Carefusion, USA. For each trait, samples with measurements that were more than 6 standard deviations away from the sample average were excluded.

Altogether, these cohorts had a total sample size of 132,200. In conducting our metanalysis, we excluded genetic variants with minor allele frequency < 0.01. We used FUMA v. 1.5.2 to annotate and functionally map variants in the meta-analysis. Genome-wide significance was defined using a threshold of P<5×10^-8^. We defined variants in the risk loci as variants correlated with the most significant variant at R^2^>0.6 using the 1000G Phase 3 EAS reference panel. Genome positions are reported in build hg37 for index variants. To designate a locus as previously known or potentially novel, the index variants, or the most significant variants in each locus, were at least 1 Mb in distance from a previously discovered genome-wide significant variant associated with the trait. Previously discovered variants were compiled from Shrine et al^33^.

### Comparison of pleiotropic effect analysis

We used S-LDSC to estimate heritability and genetic correlation of and between phenotypes using summary statistics of EUR populations. We then applied the linemodels package (https://github.com/mjpirinen/linemodels) to the GWAS summary statistics of three respiratory diseases (asthma, COPD, and lung cancer) and the three featured environmental and genetic risk factors (smoking status, smoking intensity (Cigarettes/Day), and FEV_1_/FVC) across three ancestry groups (AFR, EAS, and EUR) plus the corresponding meta-analysis results. We focused on comparisons for 12 pairs of traits selected from above. Three of these were between diseases phenotypes: asthma and COPD, asthma and lung cancer, COPD and lung cancer, and nine pairs were between disease phenotype and smoking or lung function. For each pairwise comparison, we considered variants present in the summary statistics of both traits and significantly associated with at least one of the traits being compared (**Supplementary Table 28**). We classified the variants into three classes (two when there is no variant associated with both traits) based on their association patterns: associated with trait 1 only, trait 2 only, and both. We then estimate the slopes of variants significantly associated with trait 1 and trait 2 only using an EM algorithm. Conditioning on these two classes, we ran the linemodels package on the GWAS effect sizes and standard errors of overlapped variants of the two traits, where we set the scale parameters determining the magnitude of effect sizes to 0.2, the correlation parameters determining the allowed deviation from the lines to 0.99 as default, and the slope parameter to the estimates from the previous EM step. The membership probabilities in the two classes were computed separately for each variant by assuming that the classes were equally probable a priori. This analysis was also repeated but for three classes (trait 1, trait 2, or both). We assumed no overlapping samples between the two GWASs being compared and set the correlation of their effect estimators to 0. Confident associations are defined as having a posterior probability above 0.99.

### Multi-trait analysis

We conducted multi-trait genome-wide association studies as implemented in MTAG v. 1.0.7 for each ancestry group (AFR, AMR, EAS, EUR) by combining the ancestry-specific GWAS summary statistics for GBMI asthma, GBMI COPD, GWMA lung cancer, spirometry meta-analysis, and GSCAN smoking behaviors. MTAG performs a joint analysis of GWAS results from related traits to improve the number of genetic loci identified and the predictive power of polygenic scores. For each population, we used the ancestry-specific LD reference panel from the gnomAD reference panels v2.1.1. For each ancestry-specific MTAG, we included traits with χ^2^ > 1.02.

### Study population

The *All of Us* Research Program is a longitudinal cohort study that has continuously enrolled US adults 18 years or older since May 2017. The program aims to engage in one million or more US participants and places a strong emphasis on including diverse populations that have traditionally been underrepresented in biomedical research. Details of the All of Us cohort have been previously described^60^. In summary, participants of the program opt to provide self-reported data, linked health record data, and biospecimen data to be made available for research uses. The program’s primary objective is to build a resource to help researchers understand individual differences in biological, clinical, social, and environmental determinants of health and disease to advance precision health care.

Informed consent for all the participants in the All of Us Research Program are conducted in person or through an eConsent platform. The protocol was reviewed by the Institutional Review Board (IRB) of the All of Us Research Program. Data can be accessed through the All of US Research Workbench, a secure cloud-based analytic platform. Whole genome sequencing, genotyping array variant data, variant annotations, computed ancestry, and quality reports are accessible through the Controlled Tier of the AoU. This project is registered in the All of Us program under the workspace name “PRSxtra AoU”. In our analysis, we included individuals with whole genome data in the v7 Data Release, self-reported sex, and date of birth along with additional disease-dependent filtering criteria: For COPD and lung cancer, we excluded individuals who did not self-report report smoking status. For COPD, we additionally excluded individuals with homozygous polymorphism of *SERPINA1*, which encodes for a serine protease inhibitor alpha 1 antitrypsin, as this is a known risk allele associated with COPD^62^. Participants were randomly split 70% for training and 30% for validation.

### Phenotype ascertainment

We curated clinical phenotypes from All of Us using a combination of electronic health record data, and/or self-reported personal history data from the All of Us v7 Data Release. ICD codes for each phenotype and exacerbation are detailed in **Supplementary Table 29-33**.

We define smoking status and family history based on self-reported data. Previous smokers are individuals who smoked more than 100 pack years but do not currently smoke. Individuals who have never smoked more than 100 cigarettes are considered never smokers. Family history included mother, father, and siblings with the same record of disease as the participants. No family history included those who did not explicitly self-report a family history.

### PRSxtra construction

We constructed PRSxtra in a three-layer process. Layer 1 consisted of performing multi-trait meta-analysis across related traits for each ancestry population as previously described. In layer 2, we used PRS-CSx, which leverages linkage disequilibrium across discovery samples to jointly model the genetic effects across populations via a shared continuous shrinkage prior. We used the default parameters on PRS-CSx on AFR, AMR, EAS, and EUR ancestry-specific meta-analysis results. Only HapMap3 variants—a set of 3 million variants compiled by the International HapMap Project which capture common patterns of variation in a variety of human populations—were included in calculating scores. In layer 3, we use ridge regression, as implemented by the “glmnet” R package^63^, to jointly model the 39 standardized PRS (with mean 0 and variant 1) generated in layer 2 to construct an ancestry-specific PRSxtra for the three disease phenotypes: COPD, asthma, and lung cancer. In ridge regression, we used 10-fold cross-validation and minimum lambda value to estimate the weights of each PRS. PRSxtra was validated in the held-out multi-ancestry cohort from AoU.

### PRS construction

As a baseline comparison to PRSxtra, we derived trait- and ancestry-matched PRS using PRS-CS, which uses a Bayesian regression framework to infer posterior effect sizes of SNPS. We ran PRS-CS with default parameters on the trait- and ancestry-specific GWAS summary statistic.

### Statistical analysis

For COPD, asthma, and lung cancer, we placed individuals into bins by their PRS and PRSxtra deciles. In each decile, we calculated the prevalence of disease and disease exacerbation. We calculated the risk of disease and exacerbation for each decile compared to the lowest decile of PRS and PRSxtra using logistic regression models. We evaluated the performance of predicting diseases based on PRS and PRSxtra alone, as well as in a joint multi-variable model with covariates. The baseline model included age and sex. We then subsequently added clinical risk factors (smoking, family history), and PRS or PRSxtra. We evaluated the predictive performance of each model using the area under the receiving operating curve. In the full population, we used the trait- and EUR-specific PRS as the baseline for comparison. All statistical analyses were two-sided and performed with the use of R software, version 3.5 (R Project for Statistical Computing).

## Data Availability

All data produced in the present study are available upon reasonable request to the authors.

## Contributions

Y.H. M.H.C, and A.R.M designed the study. Y.H. and W.L. processed, analyzed, and conducted statistical analysis of the data. Y.H.J., Y.W., and K.T. provided methodological and statistical advice. Y.H., W.L., D.C.Q., J.A.D., M. M, M.H.C, and A.R.M interpreted the data. A.R.M. and Y.H. obtained funding. All authors provided critical feedback and revisions for the manuscript.

## References

1. Chen, S. et al. The global economic burden of chronic obstructive pulmonary disease for 204 countries and territories in 2020–50: a health-augmented macroeconomic modelling study. Lancet Glob. Health 11, e1183–e1193 (2023).

2. Sung, H. et al. Global Cancer Statistics 2020: GLOBOCAN Estimates of Incidence and Mortality Worldwide for 36 Cancers in 185 Countries. CA. Cancer J. Clin. 71, 209–249 (2021).

3. Vos, T. et al. Global burden of 369 diseases and injuries in 204 countries and territories, 1990–2019: a systematic analysis for the Global Burden of Disease Study 2019. The Lancet **396**, 1204–1222 (2020).

4. Castro-Rodriguez, J. A. The Asthma Predictive Index: early diagnosis of asthma. Curr. Opin. Allergy Clin. Immunol. 11, 157–161 (2011).

5. He, Y. et al. Prediction and stratification of longitudinal risk for chronic obstructive pulmonary disease across smoking behaviors. Nat. Commun. 14, 8297 (2023).

6. Duffy, D. L., Martin, N. G., Battistutta, D., Hopper, J. L. & Mathews, J. D. Genetics of Asthma and Hay Fever in Australian Twins. Am. Rev. Respir. Dis. 142, 1351–1358 (1990).

7. Ingebrigtsen, T. et al. Genetic influences on chronic obstructive pulmonary disease – A twin study. Respir. Med. 104, 1890–1895 (2010).

8. Silverman, E. K. Genetics of COPD. Annu. Rev. Physiol. 82, 413–431 (2020).

9. Lichtenstein, P. et al. Environmental and Heritable Factors in the Causation of Cancer — Analyses of Cohorts of Twins from Sweden, Denmark, and Finland. N. Engl. J. Med. 343, 78–85 (2000).

10. Moll, M. et al. Chronic obstructive pulmonary disease and related phenotypes: polygenic risk scores in population-based and case-control cohorts. Lancet Respir. Med. 8, 696–708 (2020).

11. Tsuo, K. et al. Multi-ancestry meta-analysis of asthma identifies novel associations and highlights the value of increased power and diversity. Cell Genomics 2, 100212 (2022).

12. Turley, P. et al. Multi-trait analysis of genome-wide association summary statistics using MTAG. Nat. Genet. 50, 229–237 (2018).

13. Autism Spectrum Disorder Working Group of the Psychiatric Genomics Consortium et al. Identification of common genetic risk variants for autism spectrum disorder. Nat. Genet. 51, 431–444 (2019).

14. Tadros, R. et al. Shared genetic pathways contribute to risk of hypertrophic and dilated cardiomyopathies with opposite directions of effect. Nat. Genet. 53, 128–134 (2021).

15. 23andMe Research Team et al. Association studies of up to 1.2 million individuals yield new insights into the genetic etiology of tobacco and alcohol use. Nat. Genet. 51, 237–244 (2019).

16. Patel, A. P. et al. A multi-ancestry polygenic risk score improves risk prediction for coronary artery disease. Nat. Med. 29, 1793–1803 (2023).

17. Inouye, M. et al. Genomic Risk Prediction of Coronary Artery Disease in 480,000 Adults. J. Am. Coll. Cardiol. 72, 1883–1893 (2018).

18. Moll, M. et al. Polygenic risk scores identify heterogeneity in asthma and chronic obstructive pulmonary disease. J. Allergy Clin. Immunol. 152, 1423–1432 (2023).

19. Martin, A. R. et al. Clinical use of current polygenic risk scores may exacerbate health disparities. Nat. Genet. 51, 584–591 (2019).

20. The 1000 Genomes Project Consortium et al. A global reference for human genetic variation. Nature 526, 68–74 (2015).

21. Bergström, A. et al. Insights into human genetic variation and population history from 929 diverse genomes. Science 367, eaay5012 (2020).

22. Feng, Y.-C. A. et al. Taiwan Biobank: A rich biomedical research database of the Taiwanese population. Cell Genomics 2, 100197 (2022).

23. Jee, Y. H. et al. Genome-wide association studies in a large Korean cohort identify novel quantitative trait loci for 36 traits and illuminates their genetic architectures. Preprint at 10.1101/2024.05.17.24307550 (2024).

24. Jee, Y. H. et al. Cohort Profile: The Korean Cancer Prevention Study-II (KCPS-II) Biobank. Int. J. Epidemiol. 47, 385–386f (2018).

25. Yengo, L. et al. A saturated map of common genetic variants associated with human height. Nature 610, 704–712 (2022).

26. Chiou, J.-S. et al. Your height affects your health: genetic determinants and health-related outcomes in Taiwan. BMC Med. 20, 250 (2022).

27. Sohail, M. et al. Polygenic adaptation on height is overestimated due to uncorrected stratification in genome-wide association studies. eLife 8, e39702 (2019).

28. Kranzler, H. R. et al. Genome-wide association study of alcohol consumption and use disorder in 274,424 individuals from multiple populations. Nat. Commun. 10, 1499 (2019).

29. Graham, S. E. et al. The power of genetic diversity in genome-wide association studies of lipids. Nature 600, 675–679 (2021).

30. Mahajan, A. et al. Multi-ancestry genetic study of type 2 diabetes highlights the power of diverse populations for discovery and translation. Nat. Genet. 54, 560–572 (2022).

31. de Lange, K. M. et al. Genome-wide association study implicates immune activation of multiple integrin genes in inflammatory bowel disease. Nat. Genet. 49, 256–261 (2017).

32. Chen, C.-Y. et al. Analysis across Taiwan Biobank, Biobank Japan, and UK Biobank identifies hundreds of novel loci for 36 quantitative traits. Cell Genomics **3**, 100436 (2023).

33. Shrine, N. et al. Multi-ancestry genome-wide association analyses improve resolution of genes and pathways influencing lung function and chronic obstructive pulmonary disease risk. Nat. Genet. 55, 410–422 (2023).

34. Pirinen, M. linemodels: clustering effects based on linear relationships. Bioinformatics 39, btad115 (2023).

35. Ge, T. et al. Development and validation of a trans-ancestry polygenic risk score for type 2 diabetes in diverse populations. Genome Med. 14, 70 (2022).

36. Ruan, Y. et al. Improving polygenic prediction in ancestrally diverse populations. Nat. Genet. 54, 573–580 (2022).

37. Ramalho, S. H. R. & Shah, A. M. Lung function and cardiovascular disease: A link. Trends Cardiovasc. Med. 31, 93–98 (2021).

38. I., A. A. E.-A., Hamdy, G., Amin, M. & Rashad, A. Pulmonary function changes in diabetic lung. Egypt. J. Chest Dis. Tuberc. 62, 513–517 (2013).

39. Lu, D.-G., Ji, X.-Q., Liu, X., Li, H.-J. & Zhang, C.-Q. Pulmonary manifestations of Crohn’s disease. World J. Gastroenterol. 20, 133–141 (2014).

40. Çolak, Y., Nordestgaard, B. G. & Afzal, S. Morbidity and mortality in carriers of the cystic fibrosis mutation *CFTR* Phe508del in the general population. Eur. Respir. J. 56, 2000558 (2020).

41. Pereira, S. V.-N., Ribeiro, J. D., Ribeiro, A. F., Bertuzzo, C. S. & Marson, F. A. L. Novel, rare and common pathogenic variants in the CFTR gene screened by high-throughput sequencing technology and predicted by in silico tools. Sci. Rep. 9, 6234 (2019).

42. Wang, Y. et al. Rare variants of large effect in BRCA2 and CHEK2 affect risk of lung cancer. Nat. Genet. 46, 736–741 (2014).

43. Stanley, S. E., Merck, S. J. & Armanios, M. Telomerase and the Genetics of Emphysema Susceptibility. Implications for Pathogenesis Paradigms and Patient Care. Ann. Am. Thorac. Soc. 13 **Suppl 5**, S447–S451 (2016).

44. Ding, Y., et al. *TERT* gene polymorphisms are associated with chronic obstructive pulmonary disease risk in the Chinese Li population. Mol. Genet. Genomic Med. 7, e773 (2019).

45. Xu, K. et al. Genome-wide association study of smoking trajectory and meta-analysis of smoking status in 842,000 individuals. Nat. Commun. 11, 5302 (2020).

46. Wang, S. et al. Significant associations of CHRNA2 and CHRNA6 with nicotine dependence in European American and African American populations. Hum. Genet. 133, 575–586 (2014).

47. Thorgeirsson, T. E. et al. A variant associated with nicotine dependence, lung cancer and peripheral arterial disease. Nature 452, 638–642 (2008).

48. Saccone, N. L. et al. Multiple distinct risk loci for nicotine dependence identified by dense coverage of the complete family of nicotinic receptor subunit (CHRN) genes. Am. J. Med. Genet. Part B Neuropsychiatr. Genet. Off. Publ. Int. Soc. Psychiatr. Genet. **150B**, 453–466 (2009).

49. Ni, Z. et al. CLPTM1L Is Overexpressed in Lung Cancer and Associated with Apoptosis. PLoS ONE 7, e52598 (2012).

50. Chen, X. F. et al. Multiple variants of TERT and CLPTM1L constitute risk factors for lung adenocarcinoma. Genet. Mol. Res. 11, 370–378 (2012).

51. James, M. A., Vikis, H. G., Tate, E., Rymaszewski, A. L. & You, M. CRR9/CLPTM1L Regulates Cell Survival Signaling and Is Required for Ras Transformation and Lung Tumorigenesis. Cancer Res. 74, 1116–1127 (2014).

52. Ortega, V. E. et al. The Effects of Rare *SERPINA1* Variants on Lung Function and Emphysema in SPIROMICS. Am. J. Respir. Crit. Care Med. 201, 540–554 (2020).

53. Stoller, J. K. & Aboussouan, L. S. α1-antitrypsin deficiency. The Lancet 365, 2225–2236 (2005).

54. Barr, R. G. et al. Pulmonary Disease and Age at Immigration among Hispanics. Results from the Hispanic Community Health Study/Study of Latinos. Am. J. Respir. Crit. Care Med. 193, 386–395 (2016).

55. Pino-Yanes, M. et al. Genetic ancestry influences asthma susceptibility and lung function among Latinos. J. Allergy Clin. Immunol. 135, 228–235 (2015).

56. Kachuri, L. et al. Gene expression in African Americans, Puerto Ricans and Mexican Americans reveals ancestry-specific patterns of genetic architecture. Nat. Genet. 55, 952– 963 (2023).

57. Cooke, C. R. et al. The validity of using ICD-9 codes and pharmacy records to identify patients with chronic obstructive pulmonary disease. BMC Health Serv. Res. 11, 37 (2011).

58. Buhr, R. G. et al. Reversible Airflow Obstruction Predicts Future Chronic Obstructive Pulmonary Disease Development in the SPIROMICS Cohort: An Observational Cohort Study. Am. J. Respir. Crit. Care Med. 206, 554–562 (2022).

59. COPDGene Investigators et al. Genetic loci associated with chronic obstructive pulmonary disease overlap with loci for lung function and pulmonary fibrosis. Nat. Genet. 49, 426–432 (2017).

60. The All of Us Research Program Investigators. The “All of Us” Research Program. N. Engl. J. Med. 381, 668–676 (2019).

61. He, Y. & Martin, A. R. We need more-diverse biobanks to improve behavioural genetics. *Nat*. Hum. Behav. 8, 197–200 (2023).

62. Li, X. et al. Genome-wide association study of lung function and clinical implication in heavy smokers. BMC Med. Genet. 19, 134 (2018).

63. Friedman, J., Hastie, T. & Tibshirani, R. Regularization Paths for Generalized Linear Models via Coordinate Descent. J. Stat. Softw. 33, (2010).

