## Supplementary Figures for "Multi-trait and multi-ancestry genetic analysis of comorbid lung diseases and traits improves genetic discovery and polygenic risk prediction"

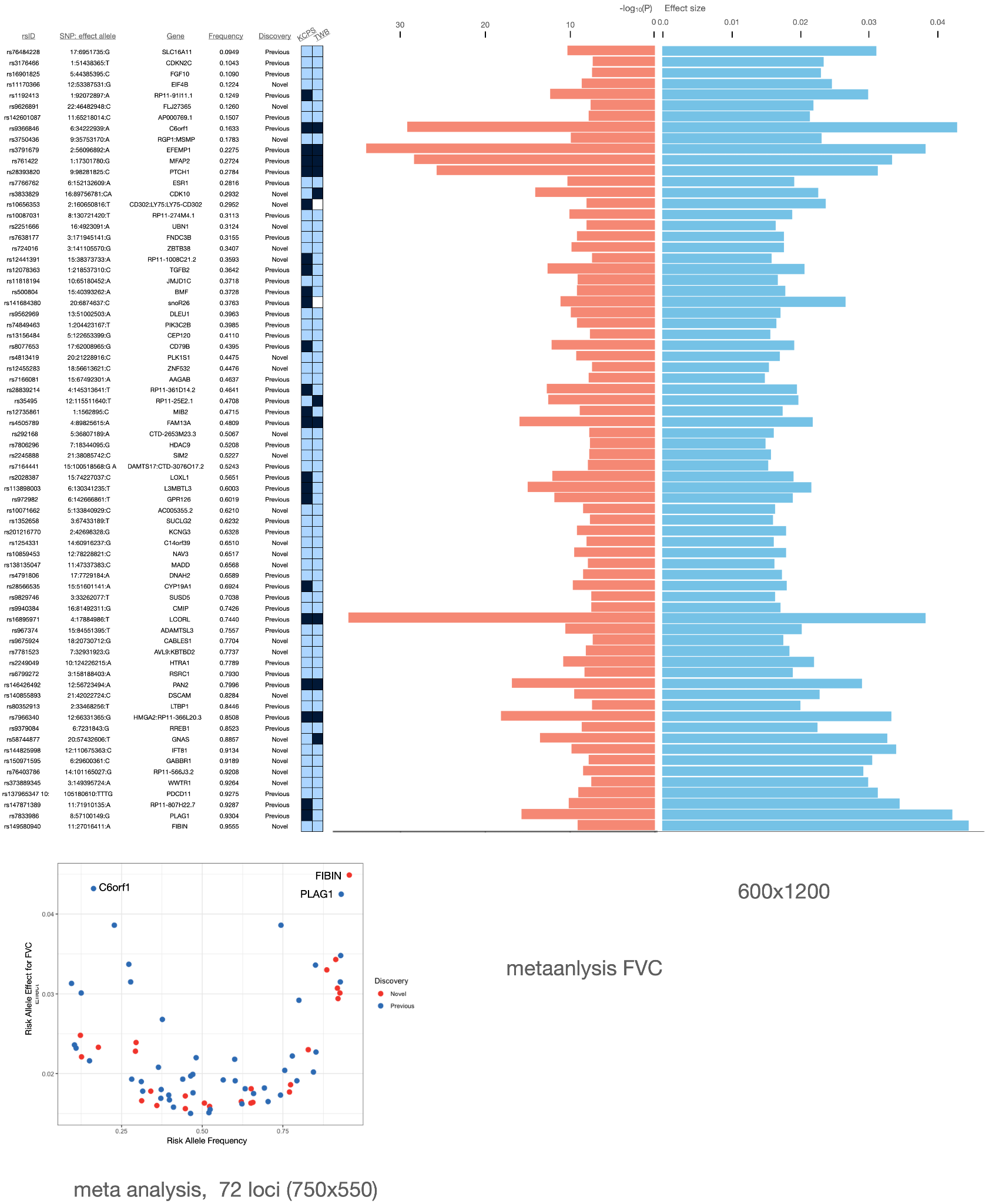

Supplementary Figure 1: Frequency and effect size of risk alleles of the 73 index variants associated with FVC in meta-analyzed GWAS of East Asian ancestry population.

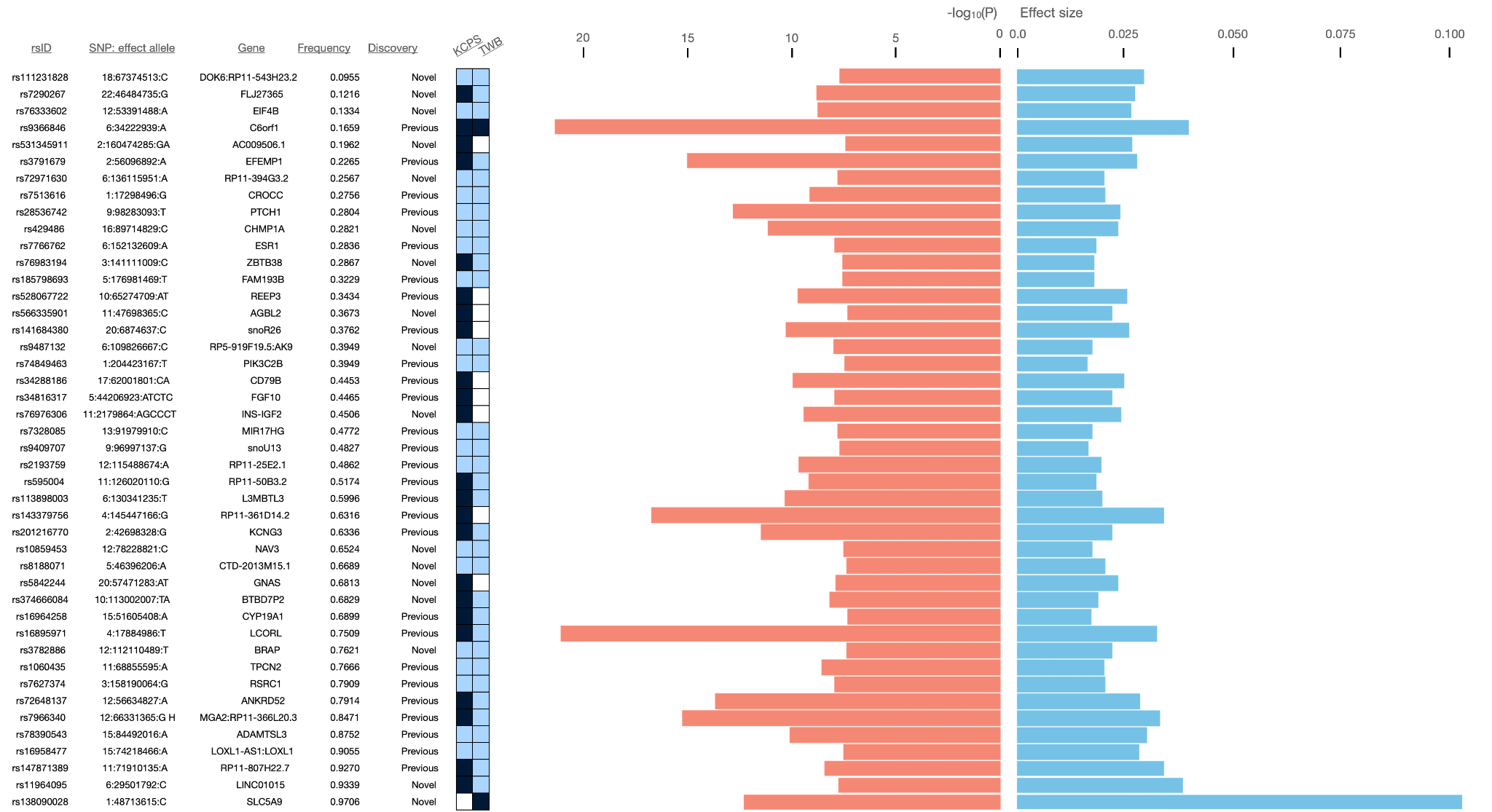

Supplementary Figure 2: Frequency and effect size of risk alleles of the 44 index variants associated with FEV_1_ in meta-analyzed GWAS of East Asian ancestry population.

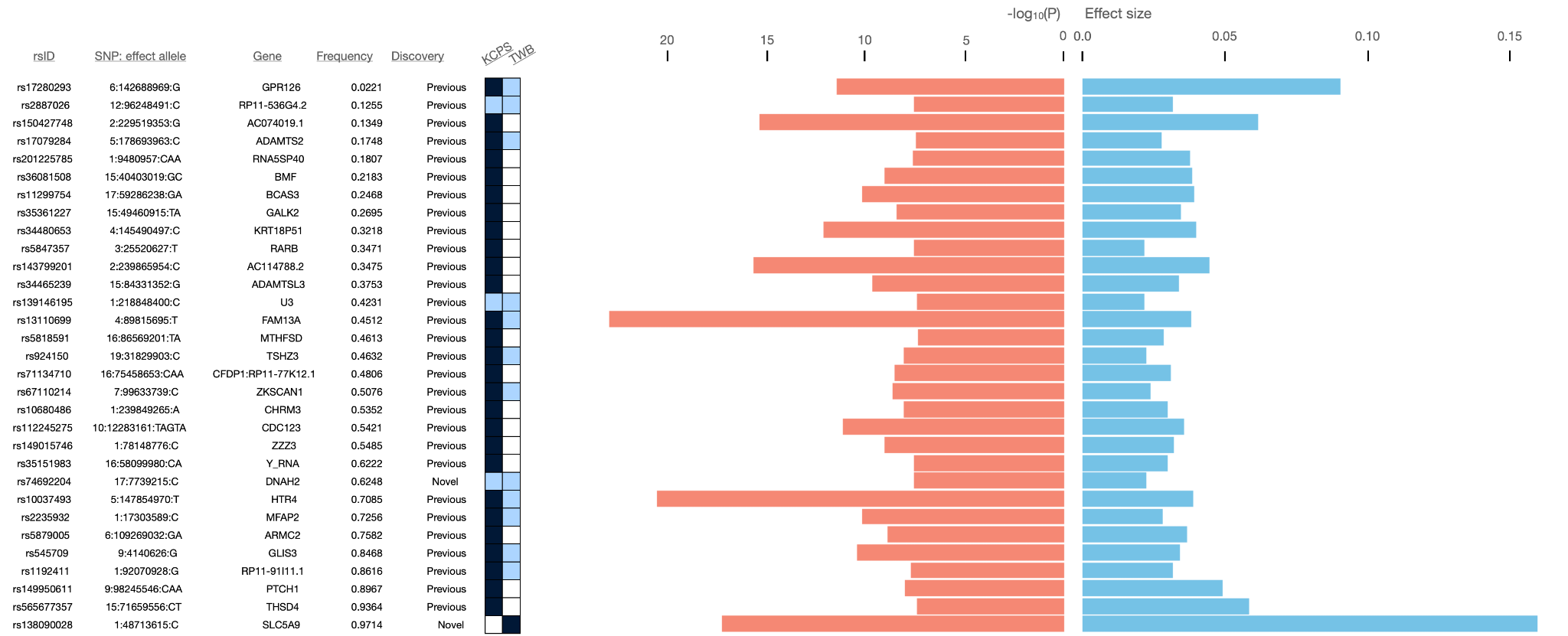

Supplementary Figure 3: Frequency and effect size of risk alleles of the 31 index variants associated with FEV_1_/FVC in meta-analyzed GWAS of East Asian ancestry population.

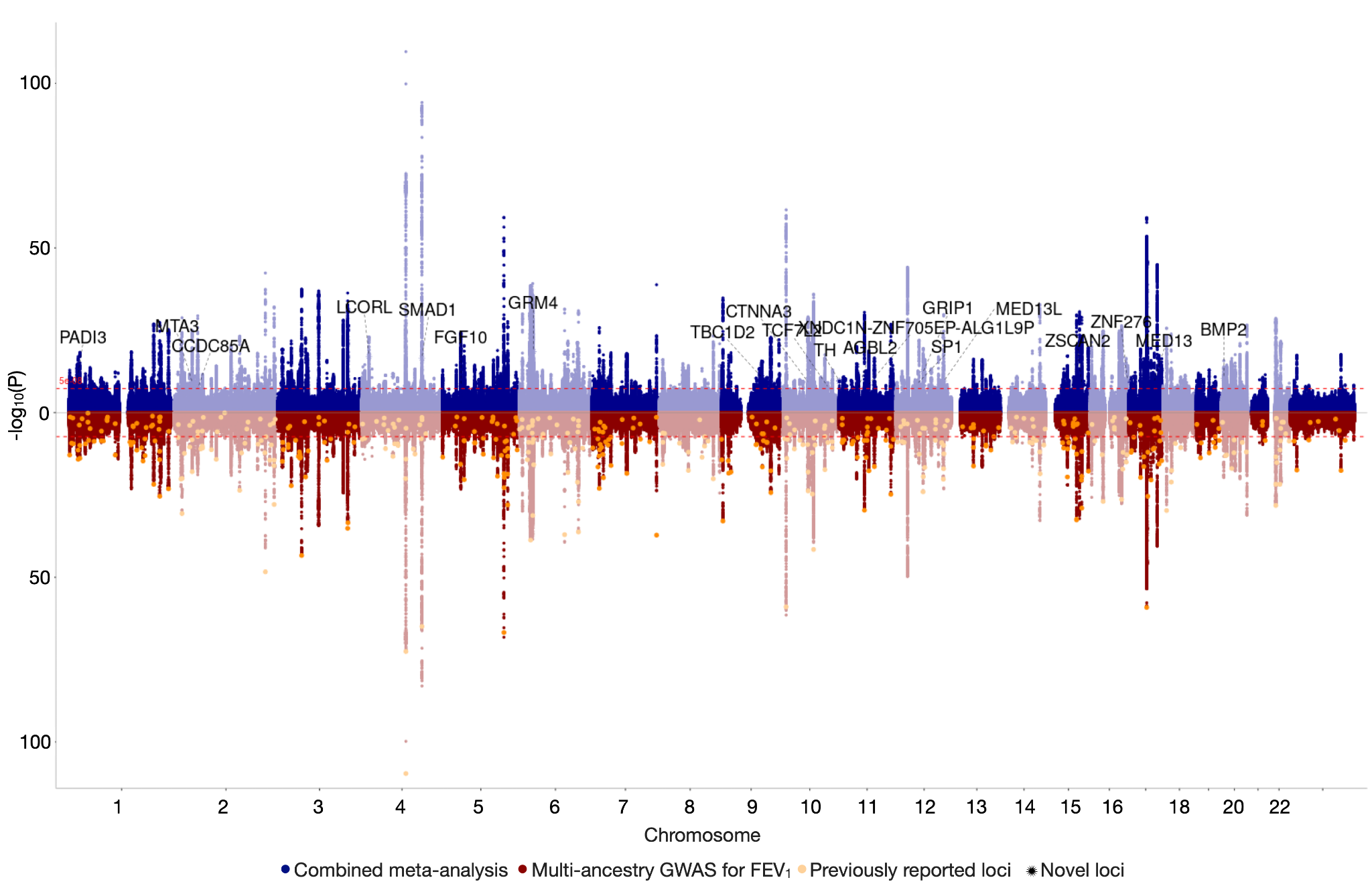

Supplementary Figure 4: Results of our combined EAS meta-analysis of FEV_1_ with the largest multi-ancestry GWAS to date. Manhattan plot of the previous multi-ancestry GWAS are shown in red, with yellow dots representing previously identified loci associated with FEV_1_. Manhattan plot of the meta-analyzed results of the previous study with our new EAS meta-analysis is shown in blue, with new loci annotated and indicated with black stars.

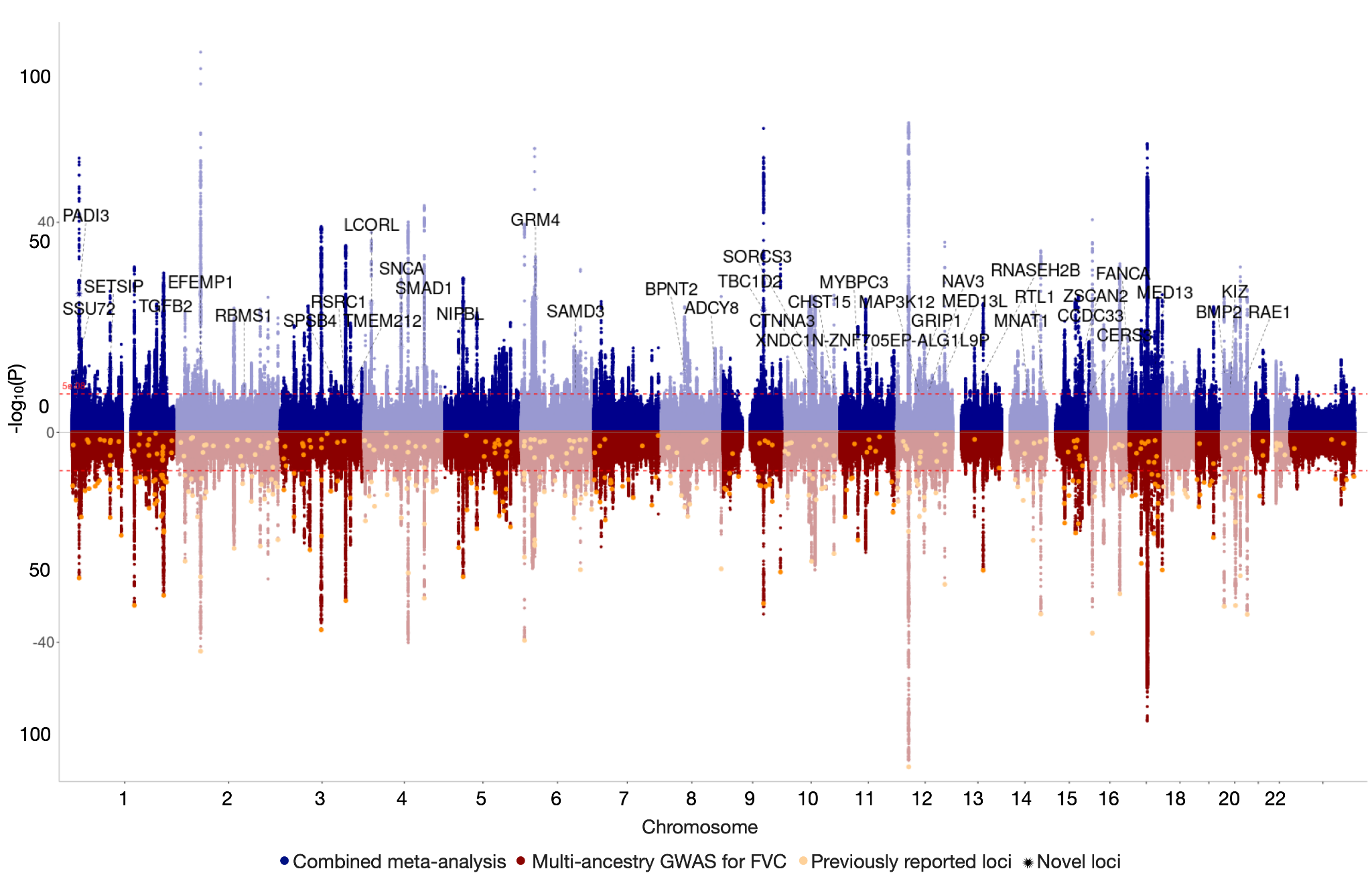

Supplementary Figure 5: Results of our combined EAS meta-analysis of FVC with the largest multi-ancestry GWAS to date. Manhattan plot of the previous multi-ancestry GWAS are shown in red, with yellow dots representing previously identified loci associated with FVCC. Manhattan plot of the meta-analyzed results of the previous study with our new EAS meta-analysis is shown in blue, with new loci annotated and indicated with black stars.

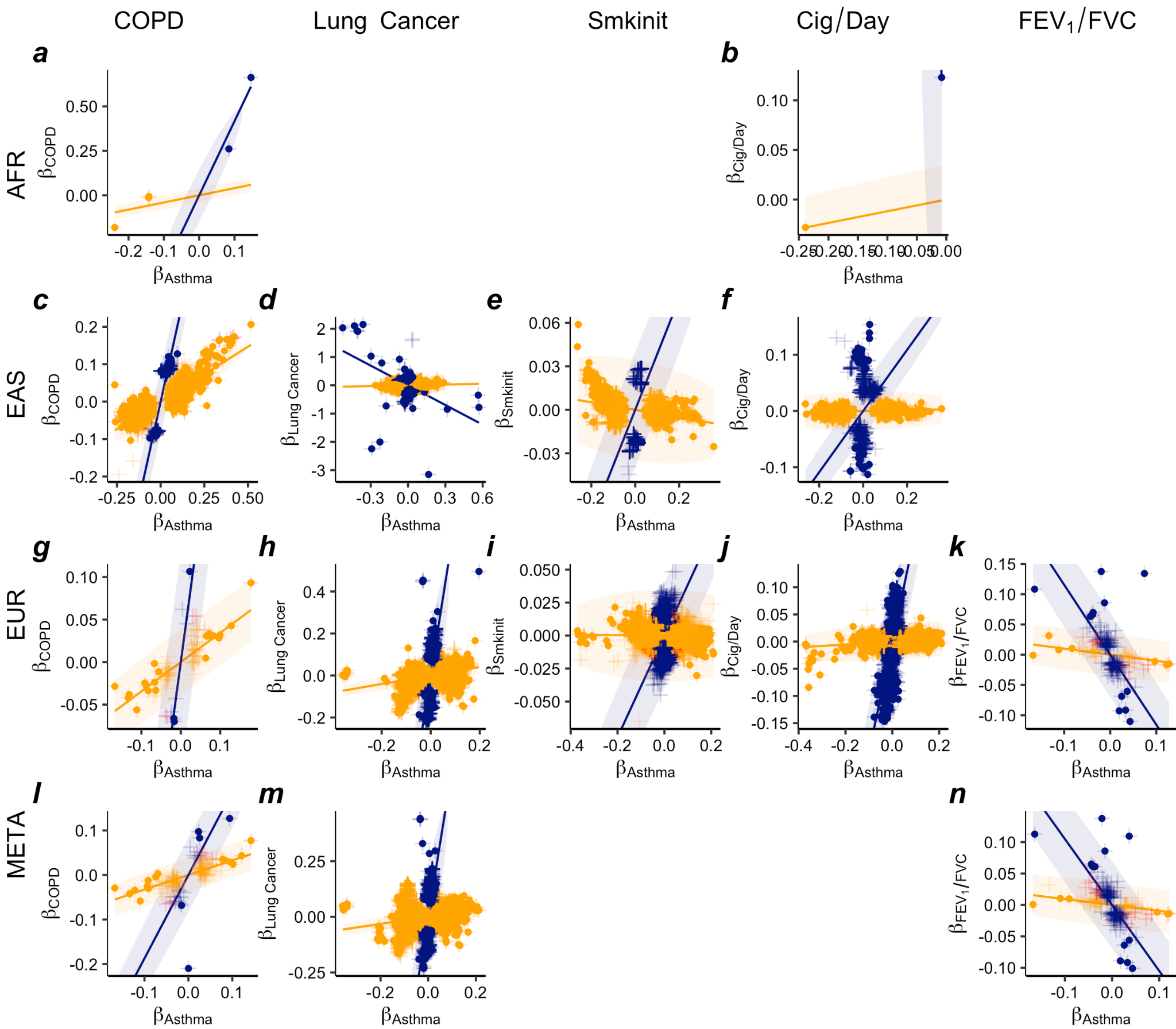

Supplementary Figure 6: Comparison of effect sizes of variants from GWAS for asthma vs. other 5 traits (columns) across all available ancestry groups (rows) in models fitted with two lines. Effect sizes of variants on asthma are on the x axis and effect sizes of variants on the other traits are on the y axis. Each cross represents a variant significantly associated (p< 5x10^-8^) with at least one of the corresponding pair of traits, colored by their significance status: variants significantly associated with both traits (red), variants significantly associated with asthma only (orange), and variants only significantly associated with the other trait (navy). Solid dots are variants confidently identified to have predominant effect on one of the two traits by a Bayesian classifier (linemodels) in shared variants analysis with posterior probability >99%: variants predominantly associated with asthma (orange), and variants predominantly associated with the other trait (navy). The colored ellipse range indicates the 95% probability regions of the fitted bivariate effect size distributions with each class. Empty space means either the two traits do not have enough overlapped variants or GWAS results are not applicable for the corresponding ancestry group.

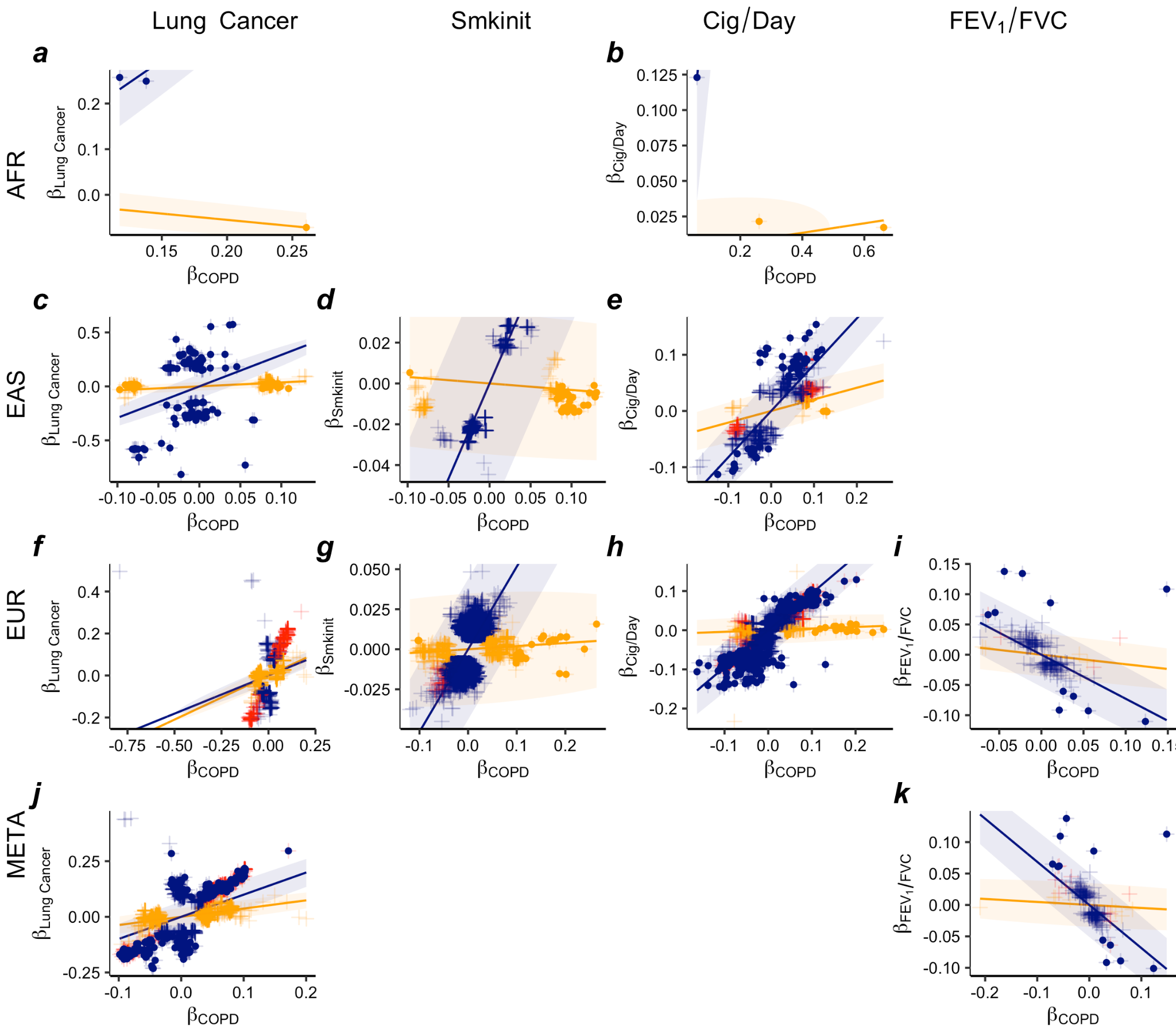

Supplementary Figure 7: Comparison of effect sizes of variants from GWAS for COPD vs. other 4 traits (columns) across all available ancestry groups (rows) in models fitted with two lines. Effect sizes of variants on COPD are on the x axis and effect sizes of variants on the other traits are on the y axis. Each cross represents a variant significantly associated (p< 5x10^-8^) with at least one of the corresponding pair of traits, colored by their significance status: variants significantly associated with both traits (red), variants significantly associated with COPD only (orange), and variants only significantly associated with the other trait (navy). Solid dots are variants confidently identified to have predominant effect on one of the two traits by a Bayesian classifier (linemodels) in shared variants analysis with posterior probability >99%: variants predominantly associated with COPD (orange), and variants predominantly associated with the other trait (navy). The colored ellipse range indicates the 95% probability regions of the fitted bivariate effect size distributions with each class. Empty space means either the two traits do not have enough overlapped variants or GWAS results are not applicable for the corresponding ancestry group.

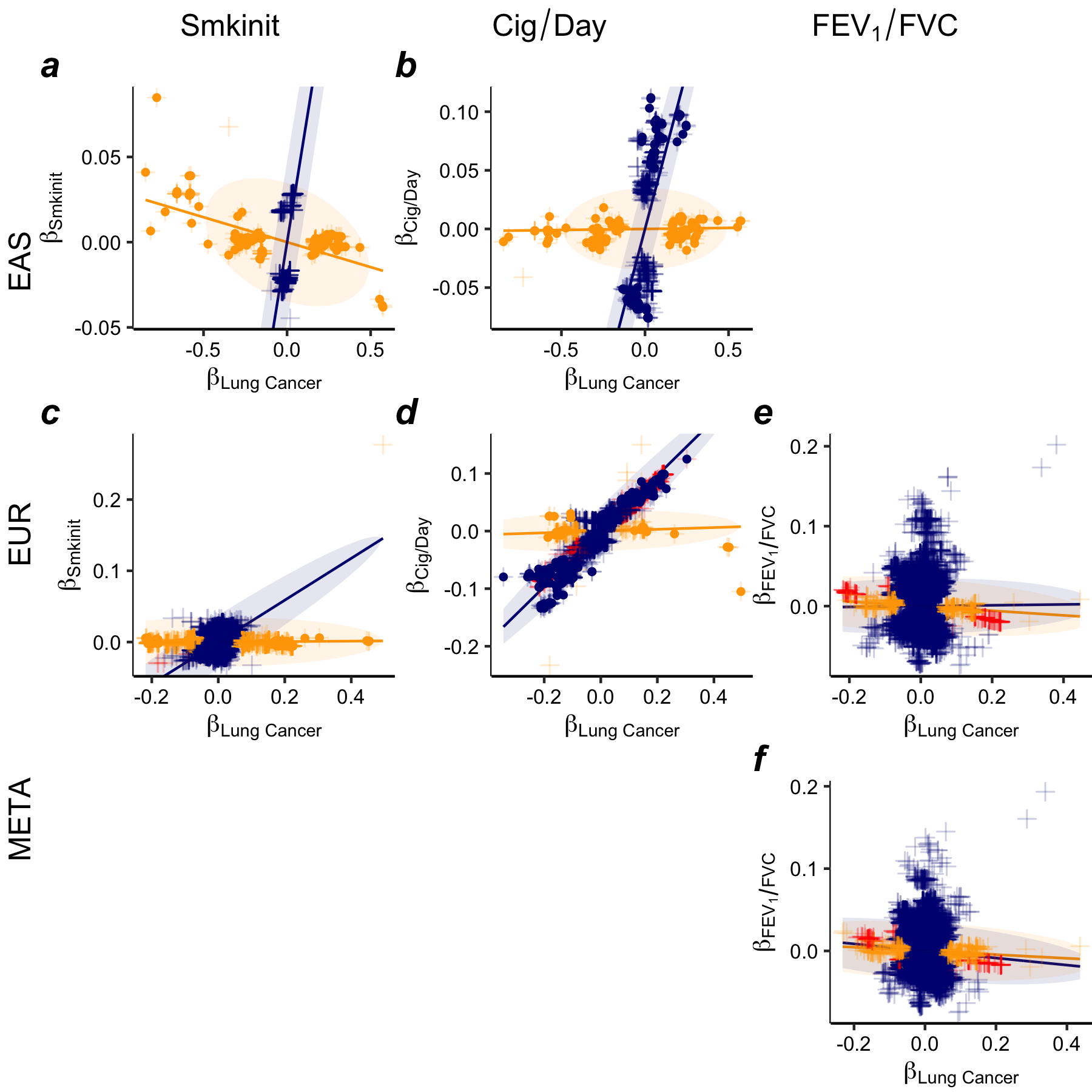

Supplementary Figure 8: Comparison of effect sizes of variants from GWAS for Lung cancer vs. other 3 traits (columns) across all available ancestry groups (rows) in models fitted with two lines. Effect sizes of variants on Lung cancer are on the x axis and effect sizes of variants on the other traits are on the y axis. Each cross represents a variant significantly associated (p< 5x10^-8^) with at least one of the corresponding pair of traits, colored by their significance status: variants significantly associated with both traits (red), variants significantly associated with lung cancer only (orange), and variants only significantly associated with the other trait (navy). Solid dots are variants confidently identified to have predominant effect on one of the two traits by a Bayesian classifier (linemodels) in shared variants analysis with posterior probability >99%: variants predominantly associated with lung cancer (orange), and variants predominantly associated with the other trait (navy). The colored ellipse range indicates the 95% probability regions of the fitted bivariate effect size distributions with each class. Empty space means either the two traits do not have enough overlapped variants or GWAS results are not applicable for the corresponding ancestry group.

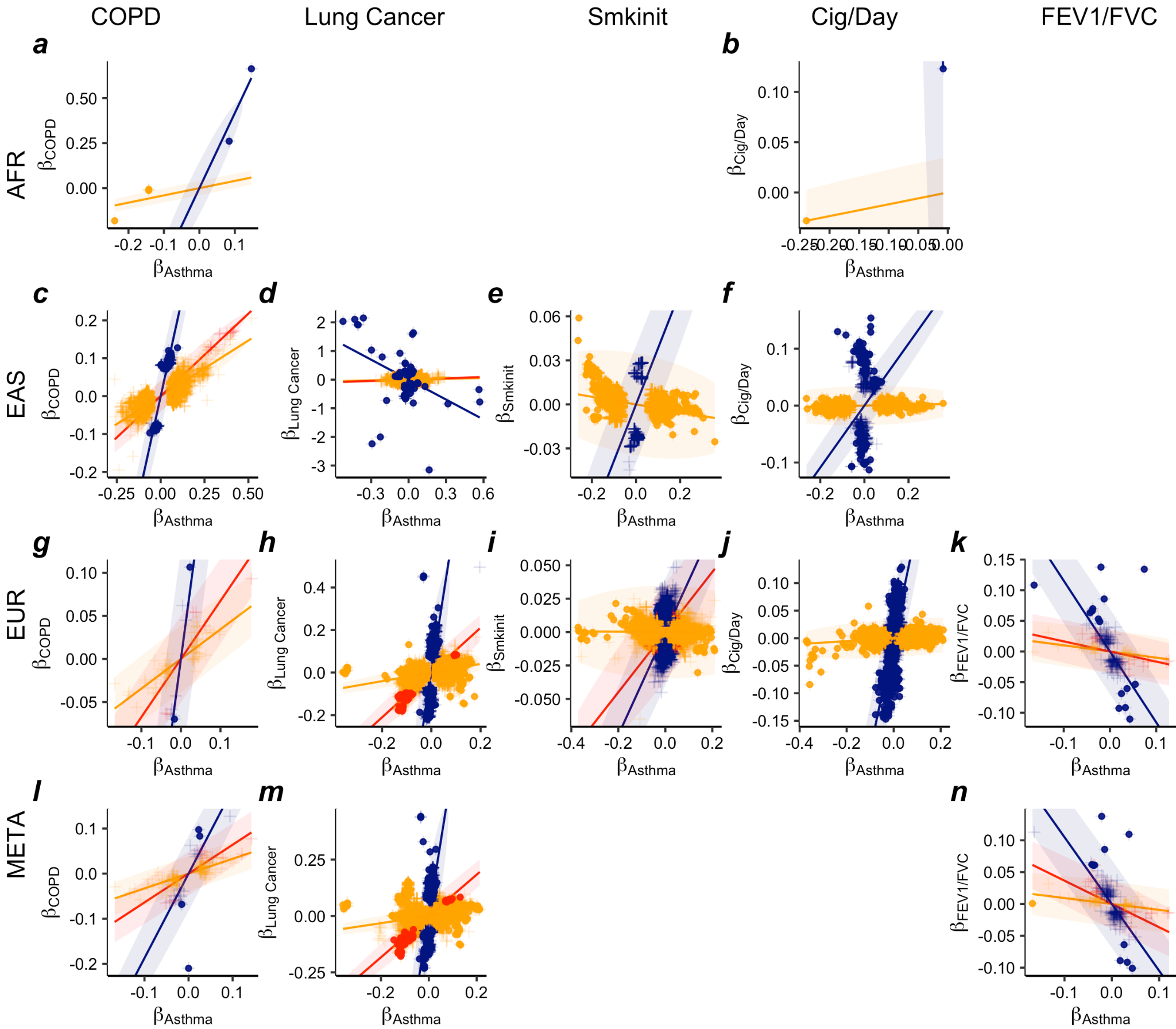

Supplementary Figure 9: Comparison of effect sizes of variants from GWAS for asthma vs. other 5 traits (columns) across all available ancestry groups (rows) in models fitted with three lines. Effect sizes of variants on asthma are on the x axis and effect sizes of variants on the other traits are on the y axis. Three distinct groups of variant effects were identified by a Bayesian classifier in shared variants analysis (colors): variants significantly associated with both traits (red), variants significantly associated with asthma only (orange), and variants only significantly associated with the other trait (navy). Solid dots are variants confidently assigned to the group indicated by the color of the dots with posterior probability >95%, while crosses are variants not confidently assigned to any of the groups. Colored ellipse range and solid line indicate the 95% probability regions of the fitted bivariate effect size distributions with each class. Empty space means either the two traits do not have enough overlapped variants or GWAS results are not applicable for the corresponding ancestry group.

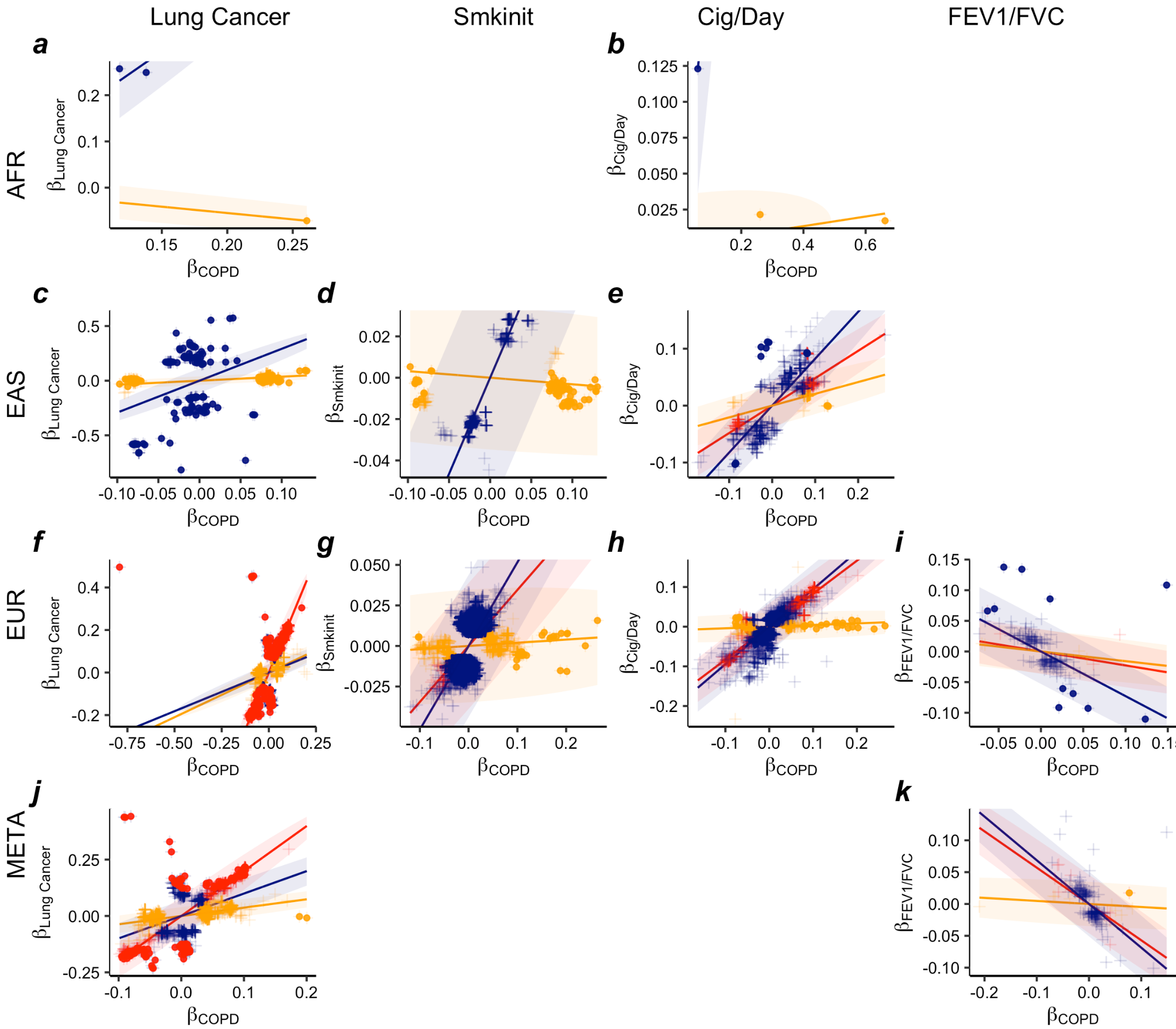

Supplementary Figure 10: Comparison of effect sizes of variants from GWAS for COPD vs. other 4 traits (columns) across all available ancestry groups (rows). Effect sizes of variants on COPD are on the x axis and effect sizes of variants on the other traits are on the y axis. Three distinct groups of variant effects were identified by a Bayesian classifier in shared variants analysis (colors): variants significantly associated with both traits (red), variants significantly associated with COPD only (orange), and variants only significantly associated with the other trait (navy). Solid dots are variants confidently assigned to the group indicated by the color of the dots with posterior probability >95%, while crosses are variants not confidently assigned to any of the groups. Colored ellipse range and solid line indicate the 95% probability regions of the fitted bivariate effect size distributions with each class. Empty space means either the two traits do not have enough overlapped variants or GWAS results are not applicable for the corresponding ancestry group.

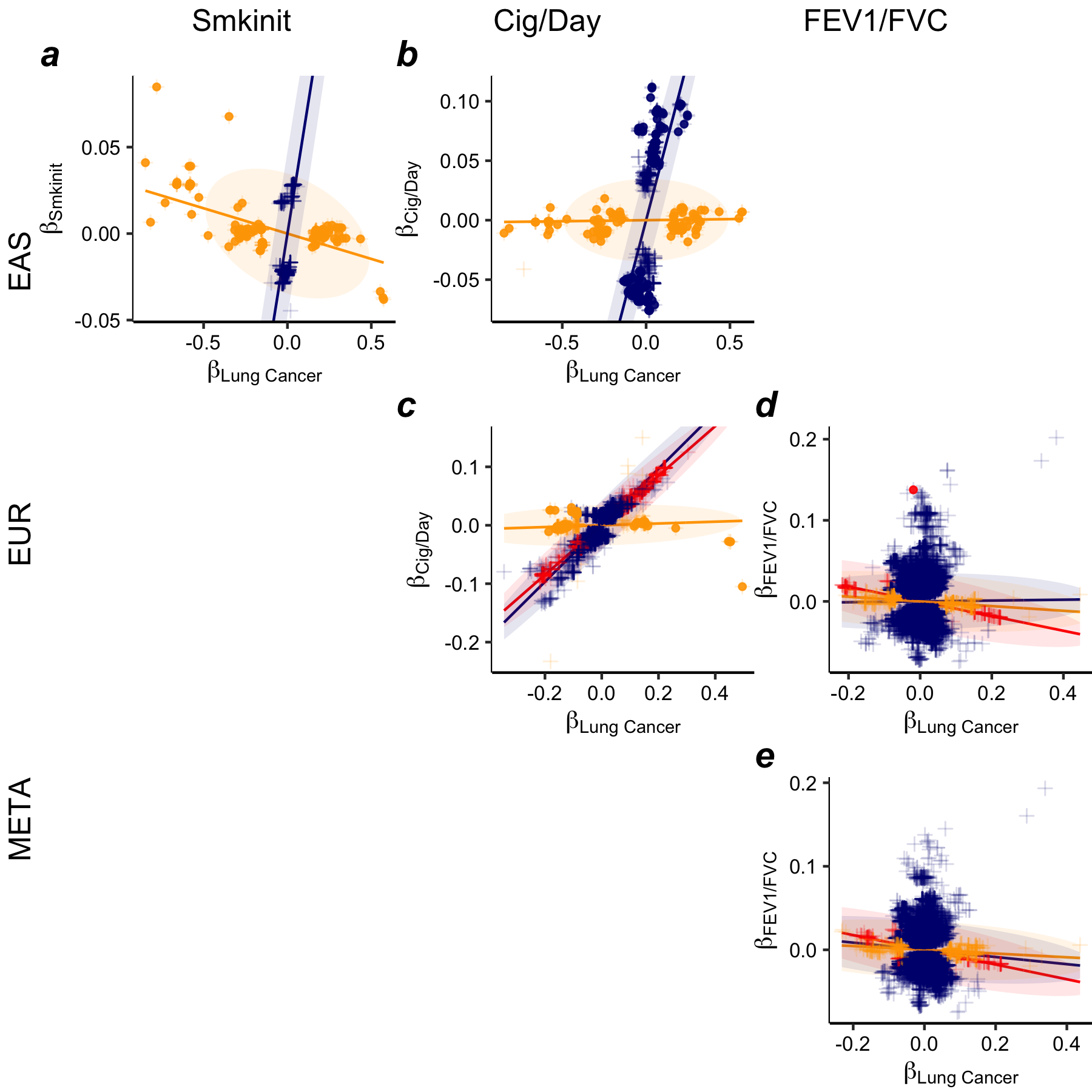

Supplementary Figure 11: Comparison of effect sizes of variants from GWAS for Lung cancer vs. other 3 traits (columns) across all available ancestry groups (rows). Effect sizes of variants on Lung cancer are on the x axis and effect sizes of variants on the other traits are on the y axis. Three distinct groups of variant effects were identified by a Bayesian classifier in shared variants analysis (colors): variants significantly associated with both traits (red), variants significantly associated with Lung cancer only (orange), and variants only significantly associated with the other trait (navy). Solid dots are variants confidently assigned to the group indicated by the color of the dots with posterior probability >95%, while crosses are variants not confidently assigned to any of the groups. Colored ellipse range and solid line indicate the 95% probability regions of the fitted bivariate effect size distributions with each class. Empty space means either the two traits do not have enough overlapped variants or GWAS results are not applicable for the corresponding ancestry group.

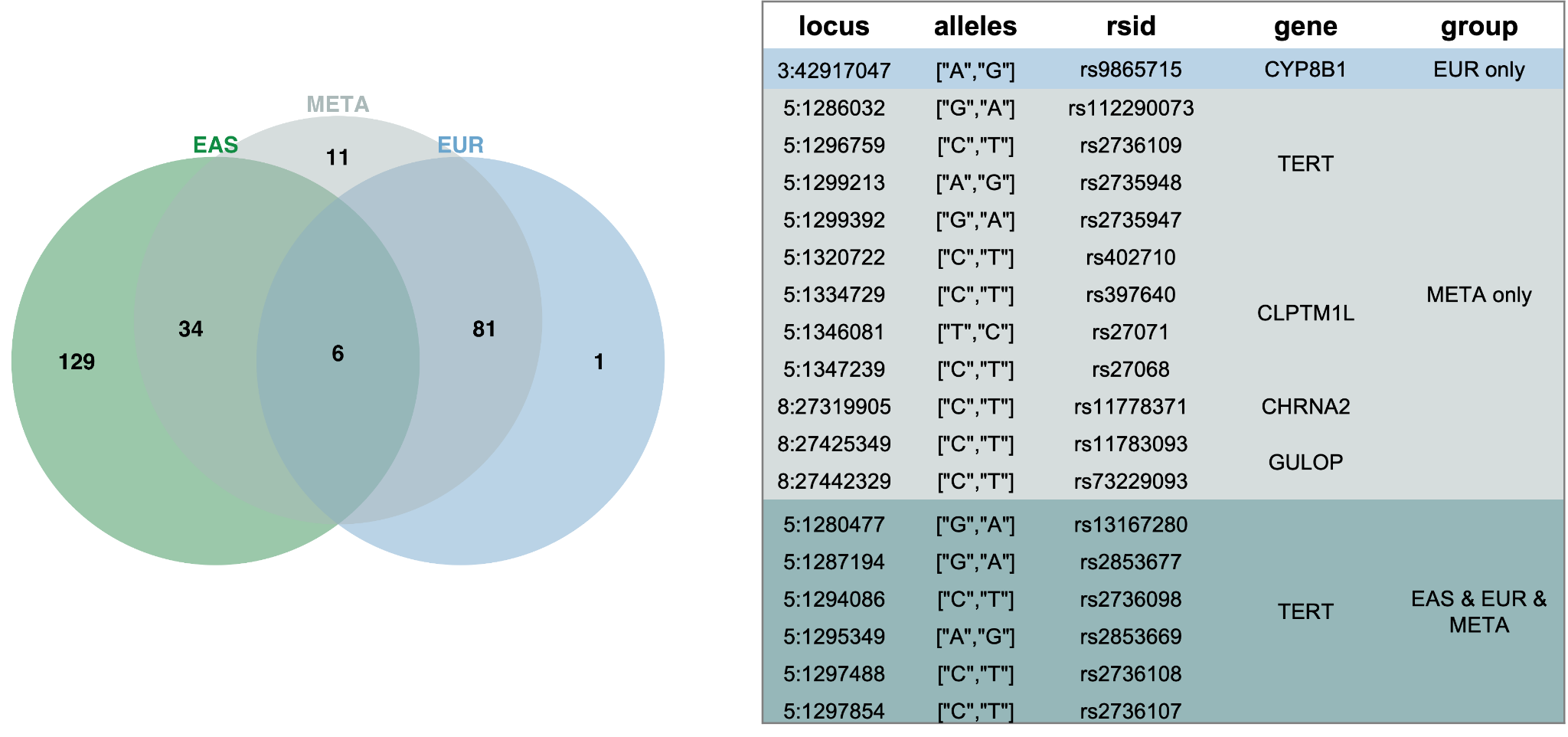

Supplementary Figure 12: Variants with lung cancer predominant effect in pleiotropy effect comparison between lung cancer and cigarette/day.

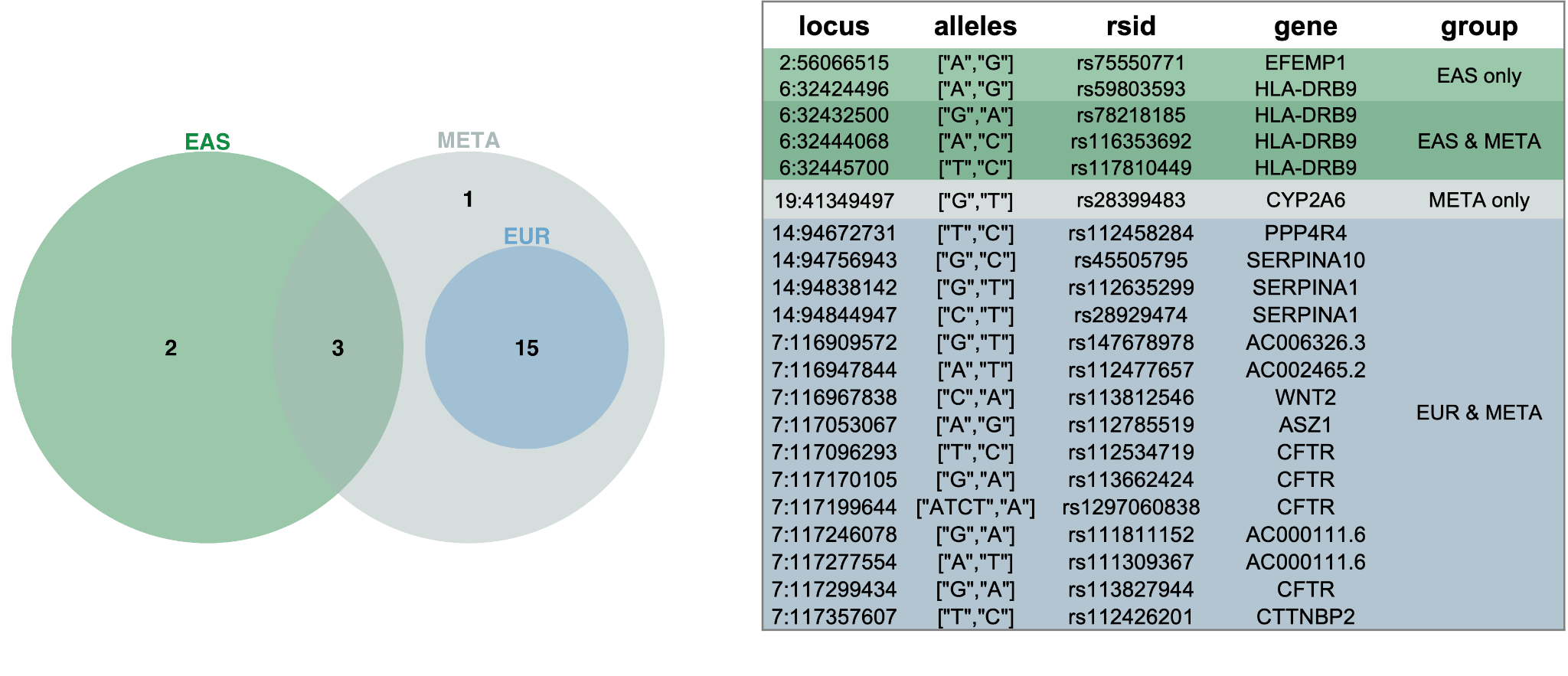

Supplementary Figure 13: Variants with COPD predominant effect in pleiotropy effect comparison between COPD and cigarette/day.
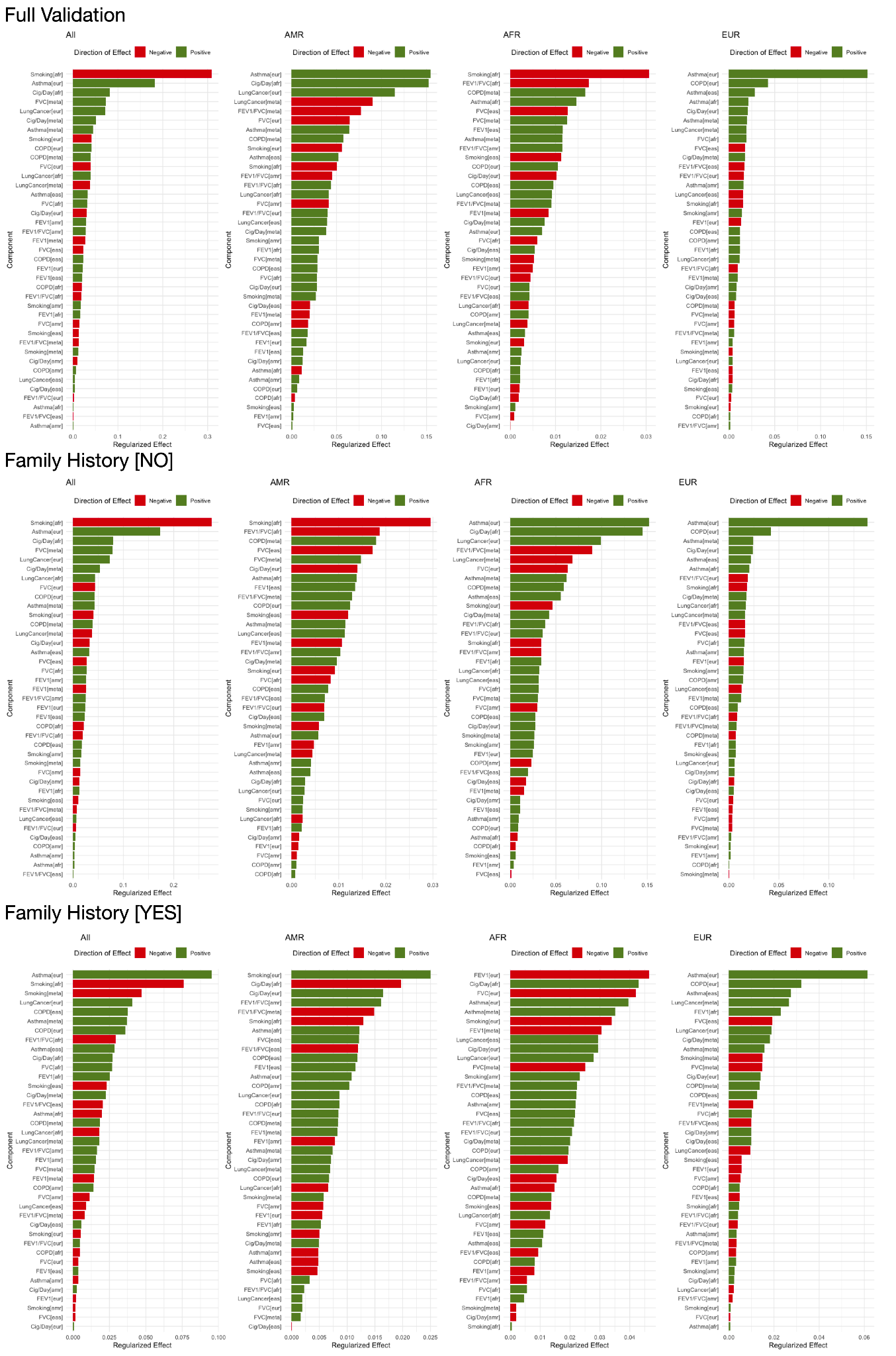

Supplementary Figure 14: Contributing weights of the components in each ancestry-specific asthma PRSxtra for the full multi-ancestry validation cohort as well as for subgroups of individuals with and without family history of asthma.

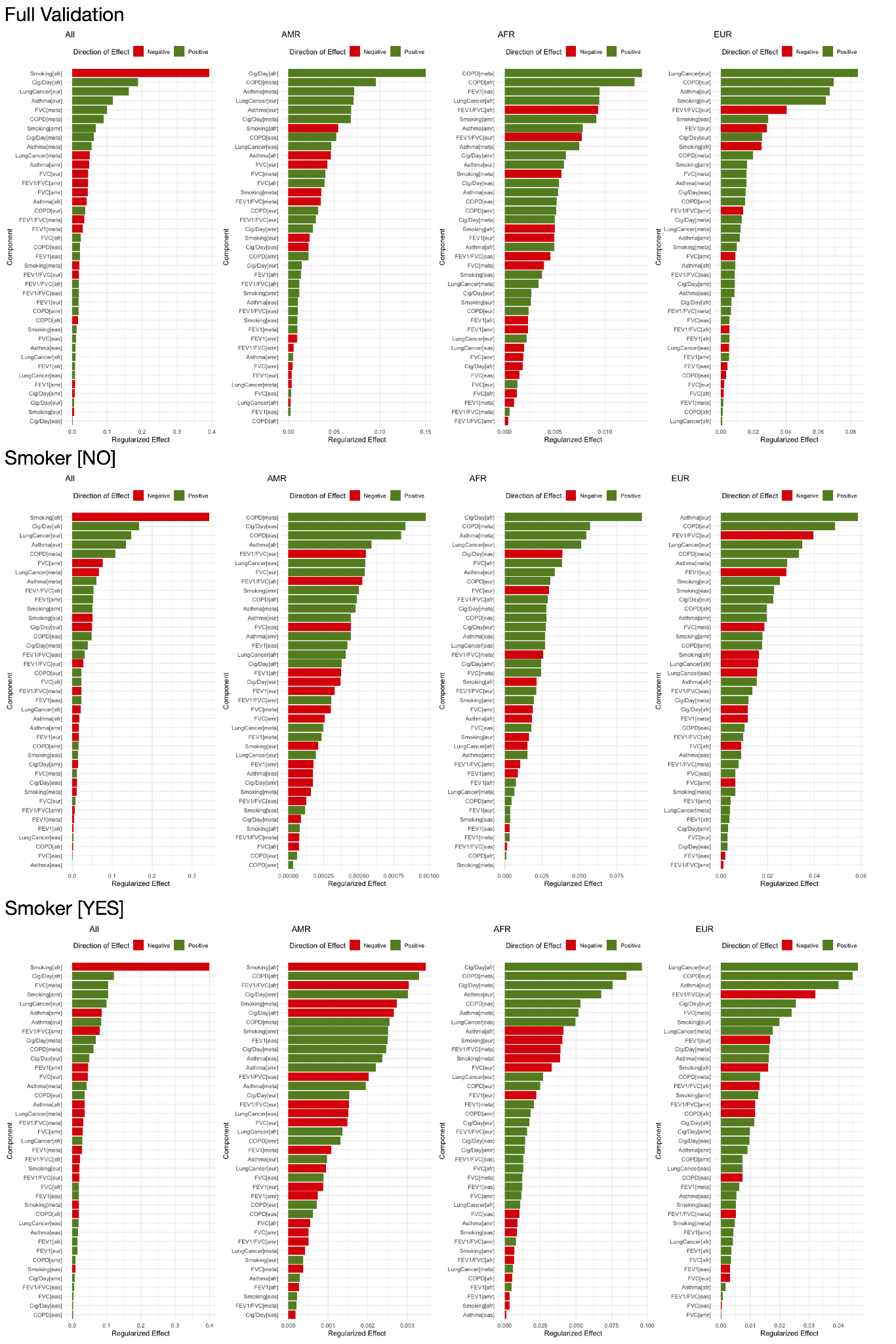

Supplementary Figure 15: Contributing weights of the components in each ancestry-specific COPD PRSxtra for the full multi-ancestry validation cohort as well as for subgroups of individuals who do and do not smoke.

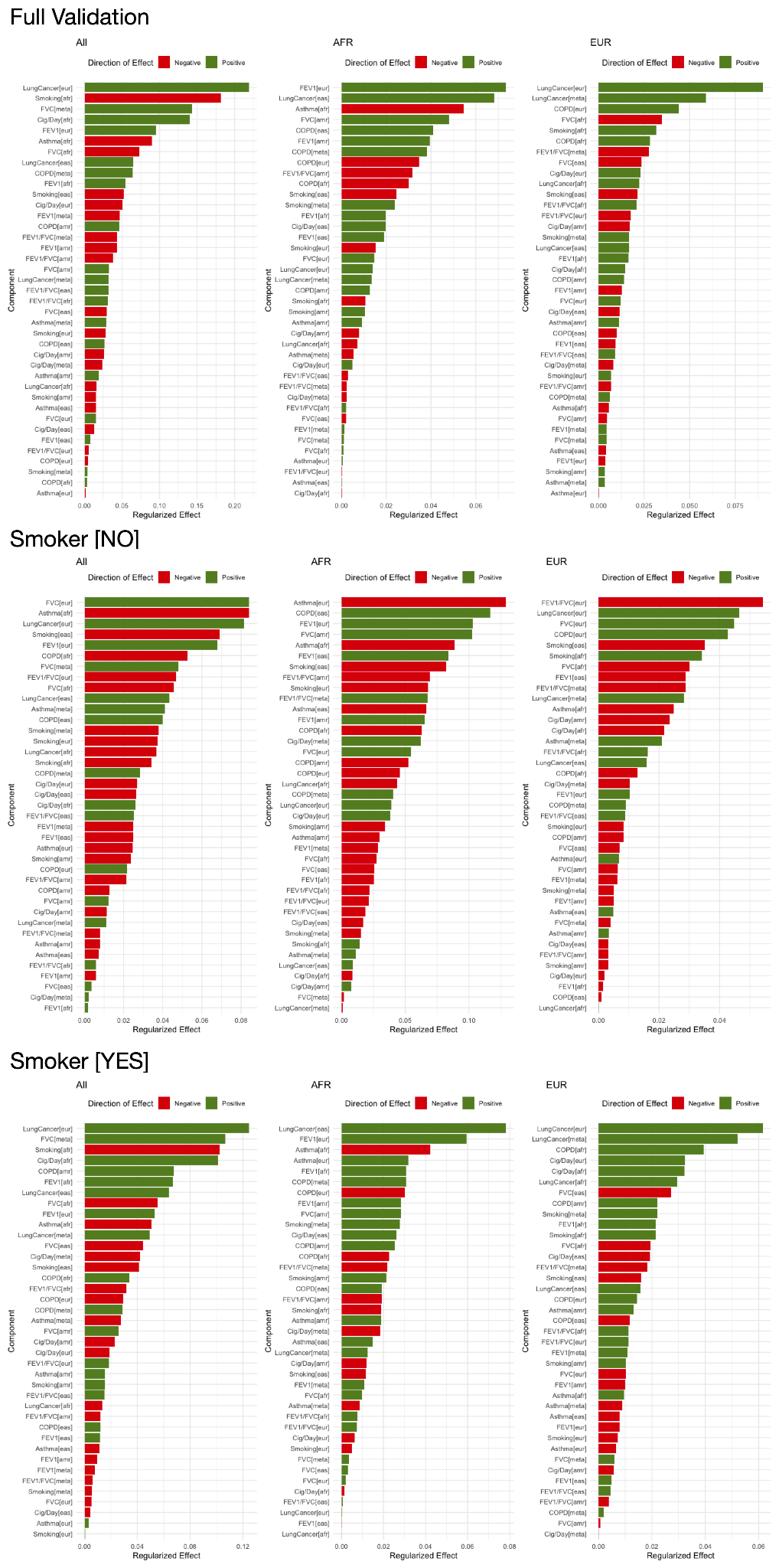

Supplementary Figure 16: Contributing weights of the components in each ancestry-specific lung cancer PRSxtra for the full multi-ancestry validation cohort as well as for subgroups of individuals who do and do not smoke.
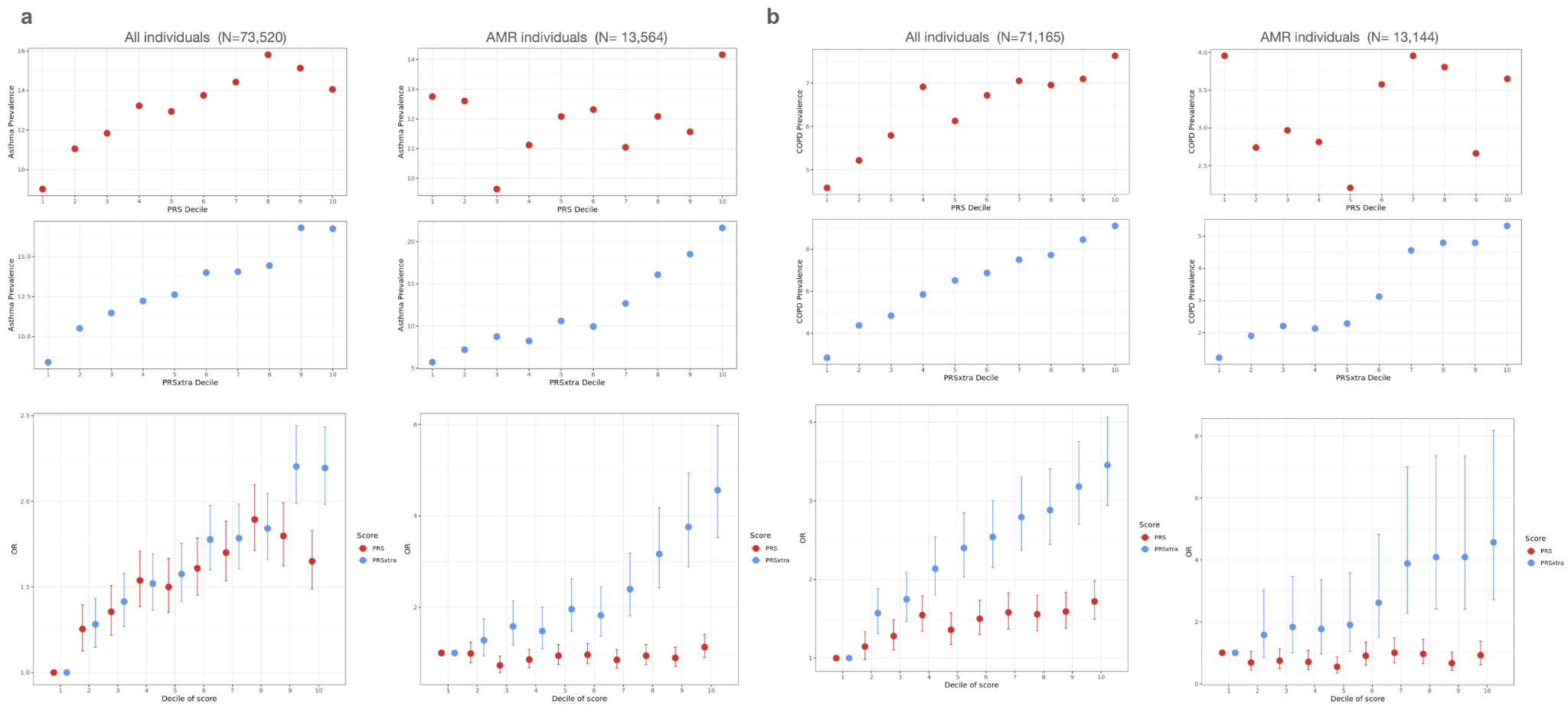

Supplementary Figure 17: Risk stratification of asthma (a) and COPD (b) by PRS and PRSxtra for in the multi-ancestry validation cohort and in AMR.

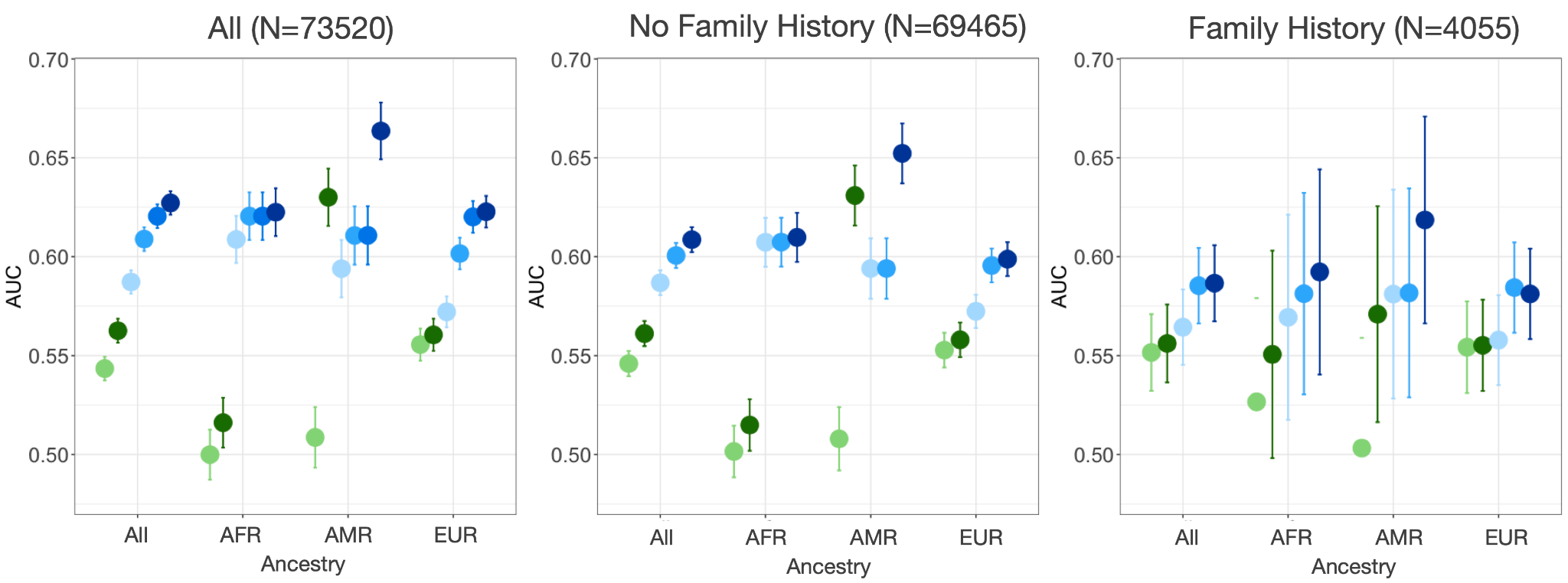

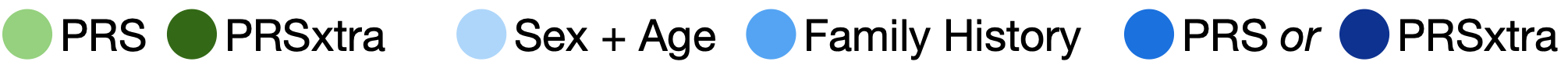

Supplementary Figure 18: Predictive power of PRS versus PRSxtra for asthma in subgroups of individuals with (right) and without (left) family history of asthma.

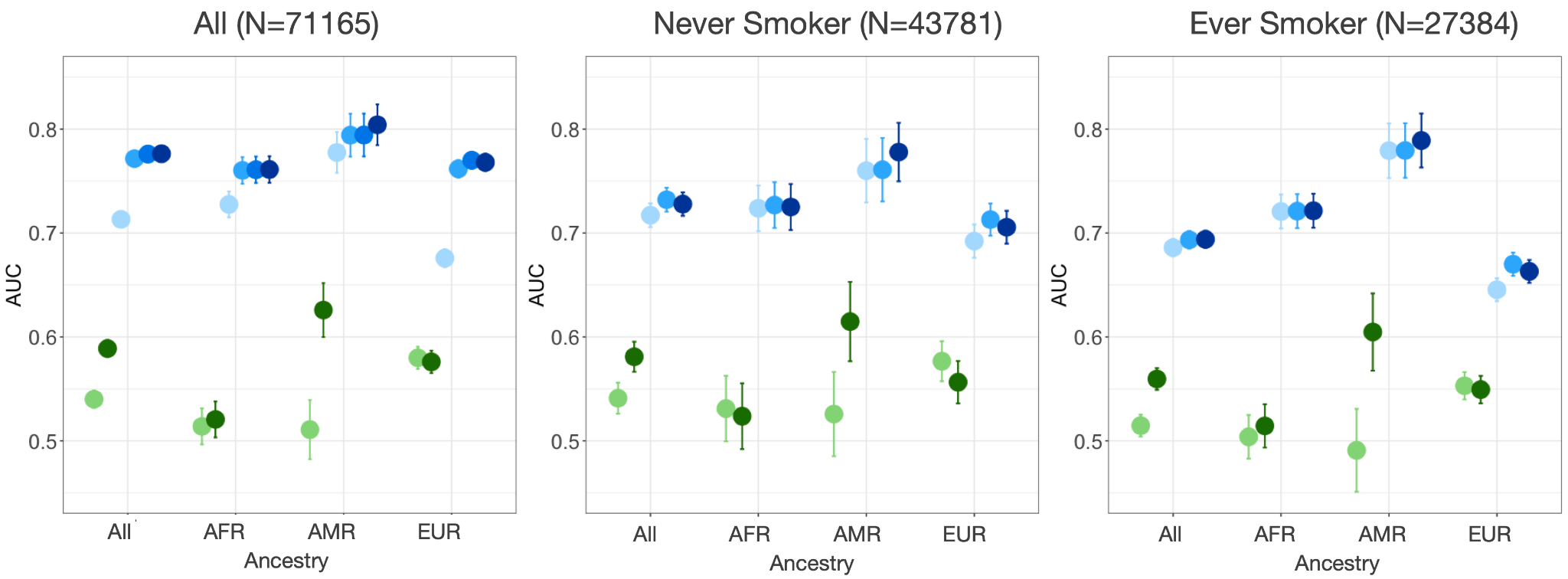

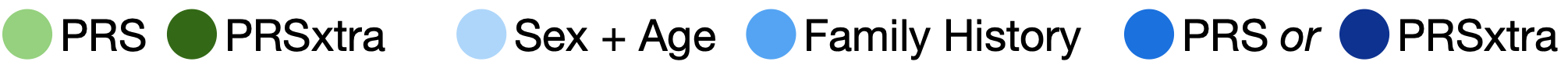

Supplementary Figure 19: Predictive power of PRS versus PRSxtra for COPD in subgroups of individuals who do (right) and do not (left) smoke.

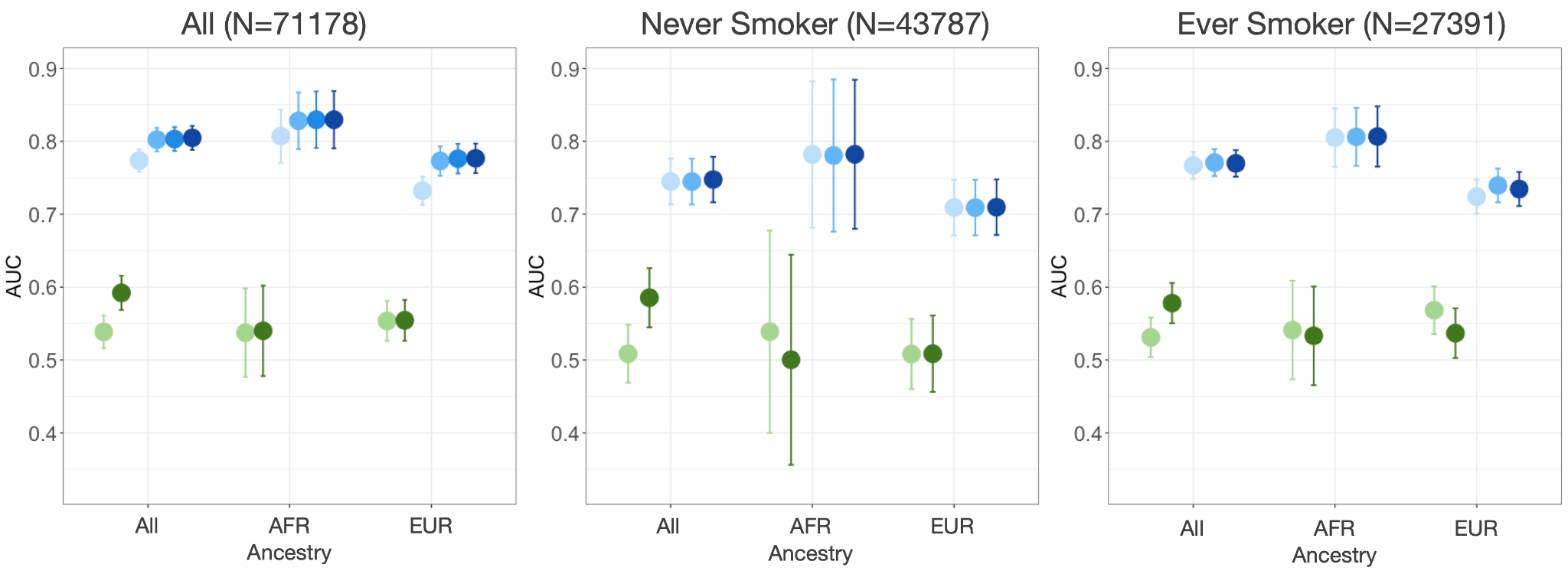

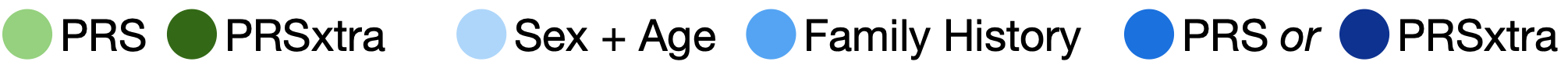

Supplementary Figure 20: Predictive power of PRS versus PRSxtra for lung cancer in subgroups of individuals who do (right) and do not (left) smoke.
